# Adolescent Cardiorespiratory Fitness and Risk of Cancer in Late Adulthood: Nationwide Sibling-Controlled Cohort Study

**DOI:** 10.1101/2024.07.01.24309761

**Authors:** Marcel Ballin, Daniel Berglind, Pontus Henriksson, Martin Neovius, Anna Nordström, Francisco B. Ortega, Elina Sillanpää, Peter Nordström, Viktor H. Ahlqvist

**Affiliations:** Department of Public Health and Caring Sciences, Clinical Geriatrics, Uppsala University, Uppsala, Sweden; Centre for Epidemiology and Community Medicine, Region Stockholm, Stockholm, Sweden; Department of Global Public Health, Karolinska Institutet, Stockholm, Sweden; Center for Wellbeing, Welfare and Happiness, Stockholm School of Economics, Stockholm, Sweden; Department of Health, Medicine and Caring Sciences, Linköping University, Linköping, Sweden; Department of Medicine, Clinical Epidemiology Division, Karolinska Institutet, Stockholm, Sweden; Department of Medical Sciences, Uppsala University, Uppsala, Sweden; School of Sports Science, UiT The Arctic University of Norway, Tromsø, Norway; Department of Physical Education and Sports, Faculty of Sport Sciences, Sport and Health University Research Institute (iMUDS), University of Granada; CIBEROBN, ISCIII, Granada, Andalucía, Spain; Faculty of Sport and Health Sciences, University of Jyväskylä, Jyväskylä, Finland; Wellbeing Services County of Central Finland, Jyväskylä, Finland; Department of Biomedicine, Aarhus University, Aarhus, Denmark; Institute of Environmental Medicine, Karolinska Institutet, Stockholm, Sweden

**Author notes:** Dr. Ballin and Dr. Ahlqvist are joint corresponding authors. Corresponding author. Prof. Nordström and Dr. Ahlqvist are joint senior authors.

## Abstract

**Objective:** To investigate whether the higher risks of certain cancers associated with high cardiorespiratory fitness can be explained by increased detection and unobserved confounders.

**Design:** Nationwide sibling-controlled cohort study of adolescents.

**Setting:** Sweden.

**Participants:** 1 124 049 men of which 477 453 were full siblings, who underwent mandatory military conscription examinations between 1972 and 1995 at a mean age of 18.3 years.

**Main outcome measures:** Hazard ratios (HR) and 95% confidence intervals (CI) of overall cancer diagnosis and cancer mortality, and 14 site-specific cancers (diagnosis or death), as recorded in the Swedish National Patient Register or Cause of Death Register until 31 December 2023, modelled using flexible parametric regressions.

**Results:** Participants were followed until a median (maximum) age of 55.9 (73.5) years, during which 98 410 were diagnosed with cancer and 16 789 had a cancer-related death (41 293 and 6908 among full siblings respectively). The most common cancers were non-melanoma skin (27 105 diagnoses & 227 deaths) and prostate cancer (24 211 diagnoses & 869 deaths). In cohort analysis, those in the highest quartile of cardiorespiratory fitness had a higher risk of prostate (adjusted HR 1.10; 95% CI: 1.05 to 1.16) and skin cancer (e.g., non-melanoma HR 1.44; 1.37 to 1.50) compared to those in the lowest quartile, which led to a higher risk of any type of cancer diagnosis (HR 1.08; 1.06 to 1.11). However, those in the highest quartile had a lower risk of cancer mortality (HR 0.71; 0.67 to 0.76). When comparing full siblings, and thereby controlling for all behavioural, environmental, and genetic factors they share, the excess risk of prostate (HR 1.01; 0.90 to 1.13) and skin cancer (e.g., non-melanoma HR 1.09; 0.99 to 1.20) attenuated to the null. In contrast, the lower risk of overall cancer mortality was still statistically significant after control for such shared confounders (HR 0.78; 0.68 to 0.89). For other site-specific cancers, the influence of such confounding tended to vary, but none showed the same excess risk as prostate and non-melanoma skin cancer.

**Conclusions:** The association between high levels of adolescent cardiorespiratory fitness and excess risk of some cancers, such as prostate and non-melanoma skin cancer, appears to be fully explained by unobserved confounders shared between full siblings. However, the protective association with cancer mortality persists even after control for such confounding.

**Summary box:** What is already known on this topic

- Adolescent physical activity and cardiorespiratory fitness are considered important factors for the prevention of cancer based on evidence from observational studies.
- Observational studies are, however, vulnerable to unobserved confounders and bias processes, including health-seeking behaviours and genetic and environmental confounders.
- These biases could explain why prior studies have found that high adolescent cardiorespiratory fitness is associated with higher risks of some cancers, typically low-mortality cancers such as prostate and non-melanoma skin cancer.

What this study adds

- This nationwide cohort study of 1.1 million male adolescents showed that while higher cardiorespiratory fitness was associated with excess risk of the most common cancers - prostate and non-melanoma skin - these associations attenuated to the null when accounting for behavioural, environmental, and genetic confounders shared between full siblings.
- In contrast, high adolescent cardiorespiratory fitness was associated with a lower risk of overall cancer mortality, which remained after controlling for unobserved confounders shared between full siblings.

## Introduction

The International Agency for Research in Cancer states that physical activity is a proven way to lower the risk of cancers,^1^ a statement also reflected through the World Health Organization guidelines for physical activity.^2^ Recently, cardiorespiratory fitness, a robust marker reflecting long-term physical activity and genetic predisposition,^3,4^ has also been implicated in cancer risk.^5–9^ Considering the challenges in identifying novel modifiable risk factors for cancer prevention,^10^ along with the downward trends in physical activity and cardiorespiratory fitness levels especially among young people,^11–14^ public health interventions targeting these traits in adolescents are gaining traction as a means of cancer prevention.

Unfortunately, the current evidence supporting the role of physical activity and cardiorespiratory fitness in cancer is associated with low certainty.^9,15^ One major limitation is that existing large-scale studies have been unable to capture important differences in behavioural, environmental and genetic confounders. This limitation is compounded by the fact that existing studies often used a composite endpoint of diagnoses and deaths, which could introduce detection bias given that individuals with high levels of physical activity more often have different health-seeking behaviours,^16^ such as higher participation in cancer screening programs.^17,18^ This increases the likelihood of early diagnosis, including for cancers typically associated with low mortality (e.g., non-melanoma skin cancer). When this is bundled together with cancer mortality, a protective effect on cancers associated with higher mortality may be underestimated or lost in the mix. Some studies have even found that higher physical activity and cardiorespiratory fitness are associated with a higher risk of overall cancer,^6,19–22^ and especially skin and prostate cancer,^6,7,18,23^ probably reflecting such bias.

These differential health behaviours remain difficult to capture and address in large-scale observational studies. One possible way to account for unobserved factors is to compare family members;^24^ who are assumed to share a large part of these behaviours and other important genetic and environmental confounders.^25,26^

In this study, we aim to address the existing inconsistencies and establish a more accurate appreciation for the cancer prevention potential of cardiorespiratory fitness. We conducted a nationwide sibling-controlled cohort study over six decades, encompassing over 1.1 million young men, of which half a million were full siblings, to investigate how the associations between adolescent cardiorespiratory and cancers in late adulthood are affected by bias processes.

## Methods

### Patient and public involvement

No patients or members of the public were involved in the study design, data collection, data analysis, interpretation of the results, decision to publish the paper or preparation of the manuscript.

### Study design

A cohort study with full-sibling analysis was conducted by cross-linking data from Swedish health and population registers, using the Personal Identification Number, which is unique for all Swedish residents.^27^ The need for informed consent was waived by the Swedish Ethical Review Authority as the study only leveraged data from existing registers.^28^ The study is reported in accordance with the STROBE guidelines.^29^

### Databases

From the Swedish Military Service Conscription Register^30^, we obtained an eligible study population based on all men who participated in military conscription examinations between 1972 and 1995, during which cardiorespiratory fitness was assessed. During this period, conscription around the age of 18 years was mandatory for all males in Sweden, with few exemptions (approximately 90% population coverage).^30^ To this, family relationships were identified using the Multi-Generation Register^31^, subsequent mortality using the National Cause of Death Register^32^, inpatient and specialist outpatient care using the National Patient Register^33^, and emigration and socioeconomic data using the registries of Statistics Sweden^31^. Overall, these data cover the entire Swedish population and are mandated by law.

### Derivation of study population

A total of 1 249 131 men were conscripted between 1972 and 1995. From these, we excluded 33 645 (2.7%) with missing data on cardiorespiratory fitness, 72 042 (5.8%) with missing data on the covariates, and 19 395 (1.5%) with extreme values (detailed below). This resulted in 1 124 049 conscripts being included in the cohort analysis (90.0% retained). Among these, 477 453 were full siblings from 219 304 families and were included in the sibling analysis.

### Exposure

Cardiorespiratory fitness was assessed at conscription using validated equipment and a standardised procedure as described in detail elsewhere.^30^ In short, conscripts performed a maximal ergometer bicycle test following normal electrocardiography. The conscript began cycling for 5 minutes at 60 to 70 revolutions per minute at a low level of predetermined (by body weight) resistance. The external resistance was then gradually increased by 25 watts every minute until exhaustion. The test results were recorded as watt maximum (W_max_), which correlates strongly with maximal oxygen uptake.^34^ Similar to in previous studies, we excluded conscripts with extremely low values (<100 W_max_).^26,35^

### Outcomes

The primary outcomes were overall cancer diagnosis and cancer mortality, as recorded in the National Patient Register and the Cause of Death register until 31 December 2023. To this, we also perform an analysis of 14 site-specific cancers (either diagnosis or death), which were selected based on their previously identified associations with physical activity or cardiorespiratory fitness:^2,5,6,19^ head and neck, oesophagus, lung, stomach, pancreas, colon, rectum, kidney, prostate, bladder, myeloma, skin (divided into melanoma and non-melanoma), and liver, bile ducts, and gallbladder. A diagnosis was ascertained as the date of first hospitalisation or visit in specialised outpatient care, recorded using diagnostic codes in a primary or secondary position, and death was classified as cancer-related if it was recorded as the primary or secondary cause on the death certificate (supplementary table 1).

### Covariates

From the Swedish Military Service Conscription Register, we collected data on age at conscription, year of conscription, and measured body mass index (BMI, kg/m^2^). As in previous studies, conscripts with extreme BMI values (≤15 and ≥60 kg/m^2^) were excluded.^26,36^ From Statistics Sweden,^37^ we obtained data on the socioeconomic status of both mothers and fathers of the conscripts, including information on the highest attained lifetime education and the annual disposable income standardised by birth years into highest-achieved quintiles between ages 40 and 50 (used to capture working life income, henceforth referred to as income categories). When values for both the mother and father were available, the highest value was retained.

### Statistical analysis

Participants were followed from the day of conscription until the date of a cancer outcome, emigration, death from non-cancer causes, or end of follow-up (31 December 2023), whichever came first. Flexible parametric survival models were used to estimate hazard ratios (HRs) for each outcome by levels of cardiorespiratory fitness, using age as the underlying time scale, and baseline knots placed at the 5^th^, 27.5^th^, 50^th^, 72.5^th^, and 95^th^ percentile of the uncensored log survival times.^38,39^ In contrast to Cox regression, these models directly estimate the baseline hazard, which enables the computation of absolute effects measures.^38^ We modelled cardiorespiratory fitness both as quartiles and using restricted cubic splines with knots placed at the 5^th^, 35^th^, 65^th^, and 95^th^ percentile.^39^ Since HRs are difficult to clinically contextualize,^40^ we also calculated the standardised cumulative incidence at 65 years of age (1-Survival) and illustrated it graphically over the follow-up period. All models were adjusted for age at conscription (continuous), year of conscription (1972, 1973-1977, 1978-1982, 1983-1987, 1988-1992, and 1993-1995), BMI (continuous and quadratic term), parental education (compulsory school ≤9 years, secondary education, post-secondary education <3 years, post-secondary education ≥3 years), and parental income (5 categories).

For the full-sibling analysis, the aforementioned model was extended to a marginalised between-within model with robust standard errors, which enables control for all unobserved confounders (including genetic and shared environmental factors).^24,41,42^ The between-within model isolates the individual-level variation (within effect) from the family-level variation (between effect) by including a term for the exposure/covariate and a term for its family average.^24,41,42^

To further aid public health interpretation, population-attributable fractions (PAF) were calculated,^43^ considering a “major” intervention that would shift everyone to the top quartile of cardiorespiratory fitness, or a “moderate” intervention that would shift everyone who belonged to the bottom quartile to the second quartile. All analyses were performed using Stata MP version 16.1.

### Sensitivity analyses

A series of sensitivity analyses were conducted to test the robustness of the results. First, to examine whether differences in estimates in sibling analysis as compared to cohort analysis were more likely to be due to selection bias rather than control for unobserved familial confounders, the standard analysis was replicated in the sibling cohort (i.e., without controlling for shared factors).^42^ Second, because BMI could be considered both a confounder or mediator in the associations between cardiorespiratory fitness and cancer,^44^ the main analysis was repeated but excluding control for BMI. Third, to examine the influence of death from non-cancer causes as a competing event, the aforementioned flexible parametric models were used to compute cause-specific cumulative incidence functions of all cancer outcomes at 65 years of age, accounting for death from non-cancer causes.^45^

## Results

### Baseline characteristics

Baseline characteristics were similar in the total cohort and in the sibling cohort (table 1). The mean (standard deviation) age at conscription was 18.3 (0.7) years and most (81.3%) were normal weight. About a quarter had parents with a high (postsecondary) level of education and about a third had parents with a high level of annual income.

**Table 1.** Baseline characteristics and number of cancer outcomes during follow-up in the total cohort and the full sibling cohort.

|  | **Total cohort**<br>**(N=1 124 049)** | **Full sibling cohort**<br>**(N=477 453)** |
| --- | --- | --- |
| **Baseline data** |  |  |
| **Birth year, median (IQR)** | 1966 (1960 to 1971) | 1965 (1961 to 1970) |
| **Age at conscription, mean (SD)** | 18.3 (0.7) | 18.3 (0.7) |
| **Body mass index, kg/m<sup>2</sup>** |  |  |
| Median (range) | 21.4 (15.0 to 59.2) | 21.3 (15.0 to 59.2) |
| **Body mass index categories, n (%)** |  |  |
| Underweight (<18.5 kg/m<sup>2</sup>) | 88 966 (7.9) | 37 884 (7.9) |
| Normal weight (18.5-24.9 kg/m<sup>2</sup>) | 913 586 (81.3) | 390 811 (81.9) |
| Overweight (25.0-29.9 kg/m<sup>2</sup>) | 102 267 (9.1) | 41 248 (8.6) |
| Obesity (>30.0 kg/m<sup>2</sup>) | 19 230 (1.7) | 7510 (1.6) |
| **Cardiorespiratory fitness, Watt max** |  |  |
| Median (range) | 271 (100 to 999) | 271 (100 to 999) |
| **Watt max by quartiles, median (range)** |  |  |
| Quartile 1 | 217 (100 to 236) | 217 (100 to 236) |
| Quartile 2 | 253 (237 to 270) | 253 (237 to 270) |
| Quartile 3 | 290 (271 to 312) | 290 (271 to 312) |
| Quartile 4 | 339 (313 to 999) | 339 (313 to 999) |
| **Parental level of education, n (%)** |  |  |
| Compulsory school <9 years | 319 242 (28.4) | 139 246 (29.1) |
| Secondary education | 505 510 (45.0) | 210 246 (44.0) |
| Post-secondary education <3 years | 123 757 (11.0) | 50 210 (10.5) |
| Post-secondary education >3 years | 175 540 (15.6) | 77 751 (16.3) |
| **Parental highest income, n (%)** |  |  |
| Category 1 (low income) | 55 055 (4.9) | 18 918 (4.0) |
| Category 2 | 108 836 (9.7) | 41 820 (8.8) |
| Category 3 | 237 953 (21.2) | 101 150 (21.2) |
| Category 4 | 338 444 (30.1) | 145 735 (30.5) |
| Category 5 (high income) | 383 761 (34.1) | 169 830 (35.6) |
| **Number of cancer events (median age at event) during follow-up** |  |  |
| Overall cancer diagnosis | 98 410 (55.1) | 41 293 (54.8) |
| Overall cancer mortality | 16 789 (55.7) | 6908 (55.3) |
| Head and neck | 4026 (54.1) | 1692 (53.7) |
| Oesophagus | 1178 (57.1) | 464 (57.0) |
| Lung | 3131 (57.4) | 1263 (57.1) |
| Stomach | 1430 (55.4) | 599 (55.3) |
| Pancreas | 2255 (57.2) | 943 (57.0) |
| Liver, bile ducts, and gallbladder | 2246 (57.1) | 925 (56.8) |
| Colon | 5320 (55.1) | 2177 (54.8) |
| Rectum | 3917 (55.7) | 1625 (55.6) |
| Kidney | 2741 (54.6) | 1151 (54.5) |
| Prostate | 24 225 (59.5) | 10 042 (59.1) |
| Bladder | 3490 (56.9) | 1432 (56.3) |
| Myeloma | 1460 (55.8) | 580 (55.9) |
| Melanoma skin | 10 026 (52.5) | 4216 (52.5) |
| Non-melanoma skin | 27 302 (54.5) | 11 509 (54.2) |

IQR = interquartile range. SD = standard deviation.

### Cancers during follow-up

During a median (range) follow-up of 37.7 (0.1 to 51.4) years, 98 410 (8.8%) were diagnosed with cancer and 16 789 (1.5%) died of cancer in the total cohort, and 41 293 (8.7%) and 6908 (1.5%) in the sibling cohort. The median age at first cancer diagnosis was 55.1 (19.1 to 73.0) years in the total cohort and 54.8 (19.1 to 72.6) years in the sibling cohort. The median age at cancer death was 55.7 (18.4 to 72.9) years in the total cohort and 55.3 (18.4 to 72.9) years in the sibling cohort. The most common site-specific cancers were non-melanoma skin (27 105 diagnoses and 227 deaths) and prostate cancer (24 211 diagnoses and 869 deaths) (table 1, supplementary table 2).

### Cardiorespiratory fitness and overall cancer

In cohort analysis, high cardiorespiratory fitness was associated with a higher risk of cancer diagnosis (fig 1, supplementary fig 1). Compared to the bottom quartile of cardiorespiratory fitness, the HR in the top quartile was 1.08 (1.06 to 1.11), with a 1.5 (1.0 to 1.9) percentage point difference in the standardised cumulative incidence at 65 years of age (supplementary tables 3-4). Calculations of PAFs produced negative estimates, implying a theoretical increase in the proportion of cancer diagnoses should either a moderate or major intervention be carried out (supplementary table 5). In full-sibling analysis, the association attenuated to the null (HR 1.00; 0.95 to 1.06; incidence difference 0.08 (-0.9 to 1.1) percentage points).

**Figure 1.**
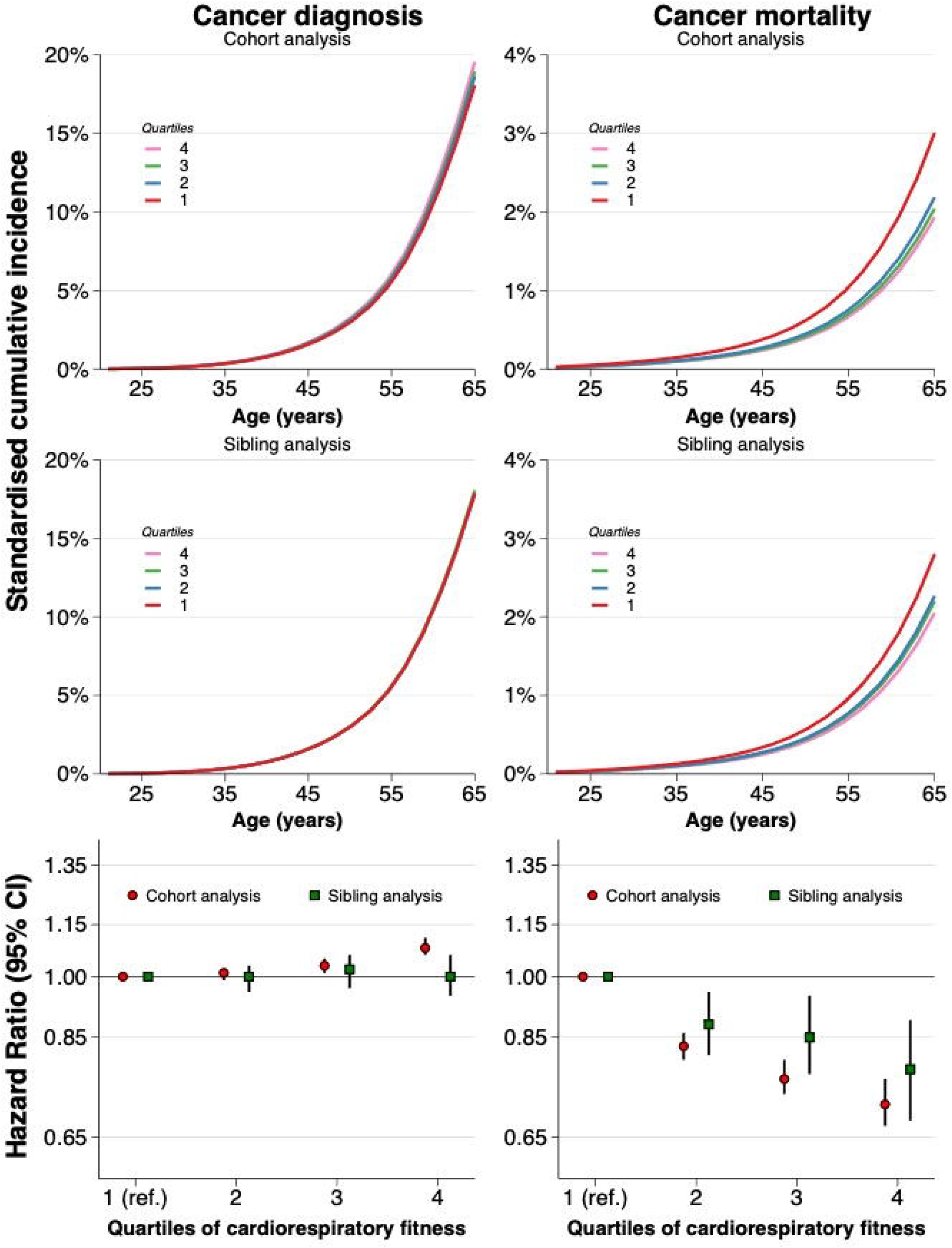
Standardised cumulative incidences and hazard ratios for overall cancer diagnosis and cancer mortality by quartiles of cardiorespiratory fitness in cohort and sibling analysis. Estimates obtained using flexible parametric survival models, extended to a marginalised between-within model in the sibling cohort, with baseline knots placed at the 5^th^, 27.5^th^, 50^th^, 72.5^th^, and 95^th^ percentile of the uncensored log survival times, and using age as the underlying time scale. The bottom quartile was the referent. All models were adjusted for age at conscription, year of conscription, body mass index, parental education, and parental income. Inferential measures for the incidences were omitted for clarity as they are also reflected by the hazard ratios and are reported in supplementary tables 3 and 4.

In cohort analysis, high cardiorespiratory fitness was associated with a lower risk of cancer mortality (fig 1, supplementary fig 1). Compared to the bottom quartile, the HR in the top quartile was 0.71 (0.67 to 0.76, with an incidence difference of -1.1 (-1.2 to -0.9) percentage points (supplementary tables 3, 4). The PAF of cancer mortality associated with a moderate intervention was 16.5% (14.4 to 18.6) and for a major intervention 22.6% (19.2 to 26.1) (supplementary table 5). In full-sibling analysis, the association only modestly attenuated (HR 0.78; 0.68 to 0.89; incidence difference -0.7 (-1.1 to -0.4) percentage points). As such, the PAF associated with a moderate intervention dropped to 11.3% (6.1 to 16.6) and for a major intervention 17.2% (8.2 to 25.5).

### Cardiorespiratory fitness and site-specific cancer

In cohort analysis, high cardiorespiratory fitness was associated with a lower risk (HRs ranging from 0.81 to 0.49) of the rectum, head and neck, bladder, stomach, pancreas, colon, kidney, liver, bile ducts, and gallbladder, oesophagus, and lung cancer (fig 2, supplementary figs 1-3, supplementary tables 3-4). In contrast, high cardio-respiratory fitness was associated with a higher risk of prostate (HR 1.10; 1.05 to 1.16; incidence difference 0.6 (percentage points), melanoma (HR 1.50; 1.45 to 1.60; incidence difference 1.0 percentage points), and non-melanoma skin cancer (HR 1.44; 1.37 to 1.50; incidence difference 2.4 percentage points).

**Figure 2.**
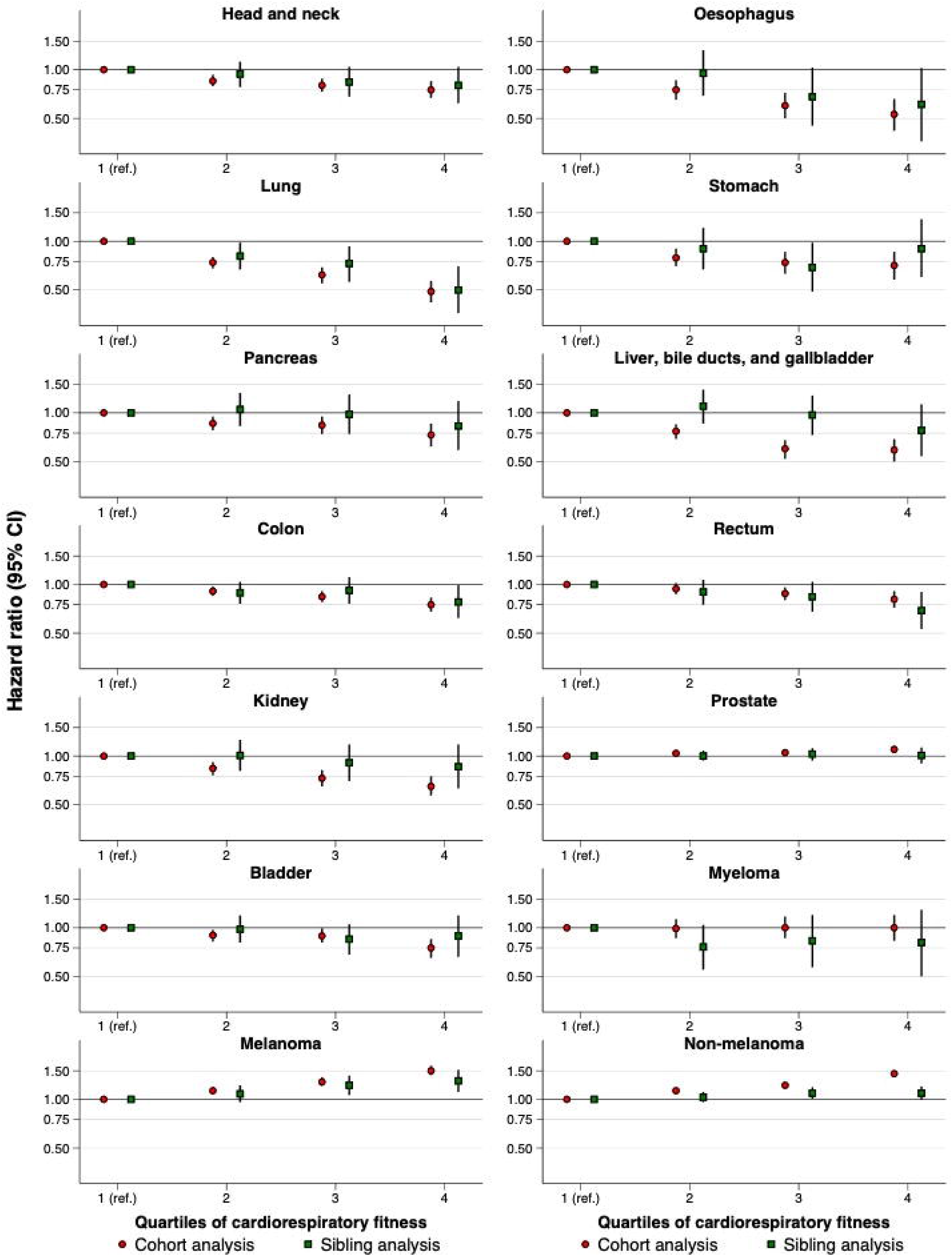
Hazard ratios for diagnosis or death from site-specific cancers by quartiles of cardiorespiratory fitness in cohort and sibling analysis. Estimates were obtained using flexible parametric survival models, extended to a marginalised between-within model in the sibling cohort, with baseline knots placed at the 5^th^, 27.5^th^, 50^th^, 72.5^th^, and 95^th^ percentile of the uncensored log survival times, and using age as the underlying time scale. The lowest quartile was the referent. All models were adjusted for age at conscription, year of conscription, body mass index, parental education, and parental income.

In full-sibling analysis, the degree of change in the associations, as compared to in the total cohort, varied between cancer types. The excess risk of non-melanoma skin cancer was largely attenuated (HR 1.09; 0.99 to 1.20; incidence difference 0.6 percentage points) as was the risk for prostate cancer (HR 1.01; 0.90 to 1.13; incidence difference 0.08 percentage points). Other tendencies toward clear attenuation were observed also for liver, bile ducts, and gallbladder cancer (from HR 0.60 to 0.97) and for kidney cancer (from HR 0.65 to 0.86), translating into reduced PAFs (supplementary table 5).

### Sensitivity analyses

Replication of the standard analysis in the sibling cohort yielded overall similar estimates as observed in the cohort analysis (supplementary table 6). The exception was rectum cancer, for which the estimates were more similar to those observed in sibling analysis. Excluding adjustment for BMI resulted in similar or slightly weaker associations for most outcomes, with a similar pattern in the cohort and sibling analysis (supplementary table 7). Accounting for the competing risk of non-cancer death (so-called crude risks) was similar to standard analysis across most outcomes (supplementary table 8). The exceptions were overall cancer diagnosis and site-specific cancers of the prostate and skin, for which the crude risks were slightly smaller.

## Discussion

### Principal findings

In this nationwide sibling-controlled cohort study spanning six decades, we find evidence suggesting that epidemiological associations between adolescent cardio-respiratory fitness and later-life cancer risk may be biased. Specifically, individuals with high adolescent cardiorespiratory fitness were at a higher risk of being diagnosed with non-melanoma skin cancer and prostate cancer. However, when adjusting for unobserved behavioural, environmental, and genetic confounders shared between full siblings, these excess risks disappeared. In contrast, high cardiorespiratory fitness was associated with lower cancer-related mortality, which remained after adjustment for such confounding.

### Comparisons with other studies

Physical activity and cardiorespiratory fitness have been linked to lower cancer risk, although the certainty in the evidence is limited, including a largely unknown contribution of unobserved confounding and bias processes.^5–9,19,25^ In this study, high adolescent cardiorespiratory fitness was linked to excess risk of non-melanoma skin and prostate cancer, as in previous observational studies and meta-analyses in men.^6,19^ However, we provide evidence to suggest that this risk appears to be fully explained by bias as it could not be replicated after accounting for unobserved confounders shared between full siblings. The sibling analysis cannot pinpoint the exact confounding factor(s), but it may be a combination of differential health-seeking behaviours (including participation in cancer screening), as well as other shared environmental factors and genetics. Previous studies showing associations between physical activity and cardiorespiratory fitness and excess risk of skin and prostate cancer have often suggested that their findings may be the result of bias,^6,18,46^ speculating that it may be explained by fit people also spending more time outdoors in the sun and that they participate in screening programs to a greater extent.^47,48^ Formal validation of these confounding and bias processes has, however, been lacking until now. Yet, even in this large-scale study, the excess risk of melanoma skin cancer could not be fully explained by confounders shared between full siblings. This may suggest the presence of biases beyond those shared by siblings, and we encourage future research to further scrutinize this association.

The association between high adolescent cardiorespiratory fitness and lower risk of overall cancer mortality remained statistically significant after accounting for such confounding factors (HR 0.78; 95% CI: 0.68 to 0.89). One previous study based on the same population reported a lower risk of both cancer diagnosis and cancer mortality in sibling analysis.^25^ Yet, interpretation of those estimates and comparisons to ours is difficult considering that the study relied on a younger cohort with substantially fewer cancer diagnoses and deaths (15 093 [1.3%] & 4900 [0.4%], compared to our study with 98 410 [8.8%] diagnoses & 16 789 [1.5%] deaths).^25^ Additionally, only time-specific hazard ratios were reported^25^, which are difficult to interpret as they are largely influenced by the depletion of susceptibles.^40^ Nevertheless, based on the results presented here, it appears important to distinguish a diagnosis of cancer from cancer-related mortality since collapsing the two may dilute the benefits associated with high-mortality cancers by introducing biased associations with a diagnosis of low-mortality cancers detected in screening. By extension, this could suggest that previous large-scale studies and meta-analyses of cardiorespiratory fitness or physical activity, which relied on composite endpoints, might have underestimated the protective associations.^5,6,19–22^ If this is true, and can be verified by future causal analysis, the potential of public health initiatives targeting cardiorespiratory fitness or physical activity to reduce cancer mortality might be underappreciated.^49,50^

This study also revealed a heterogeneous pattern regarding adolescent cardiorespiratory fitness and site-specific cancer after comparing full siblings. For example, the associations showing lower risks of kidney and liver, bile ducts, and gallbladder cancer seemed to attenuate more, whereas those with colon and head and neck cancer seemed more robust. These findings may suggest a more nuanced role of adolescent cardiorespiratory fitness in site-specific cancer, whereby there may be a greater preventive potential for certain types and less for others. Assuming part of the attenuation is the result of control for familial confounders, these findings align with research showing a clear variation in the contribution of genetic and shared environmental factors to the risk of different cancers.^51,52^

### Strengths and limitations

This study has several notable strengths, including its nationwide coverage, family-based analysis, standardised measures of cardiorespiratory fitness, extensive follow-up until late adulthood with virtually zero attrition, and investigation of both diagnoses of cancer in a universal healthcare system and cancer mortality.

However, some limitations should be considered. First, our study was limited to men due to historical conscription practices in Sweden. While we may believe there are limited reasons for sex-specific mechanisms linking cardiorespiratory fitness and physical activity to cancer, it is important to note that cancer prevalence varies by sex, and we were unable to examine cancers specific to females. Second, our study focuses on adolescent cardiorespiratory fitness and may provide insights into public health policies aimed at young individuals. However, it may not directly inform interventions targeted at other populations, despite evidence that cardiorespiratory fitness moderately tracks from adolescence into adulthood.^53^ Third, due to the limited number of cases of site-specific cancers, we were unable to conduct separate analyses of diagnosis and mortality. Furthermore, the small counts of some site-specific cancers can explain the confidence intervals overlapping the null in the sibling analysis, which is not necessarily evidence of the absence of effects. Fourth, while comparisons of full siblings are expected to control for several important confounders that are difficult to measure at scale, this design hinges on assumptions^42,54^ and there likely remains residual confounding (e.g., the remaining 50% of genetics not shared between full siblings and behaviours not shared by full siblings with different fitness levels). Future studies utilizing monozygotic twins or other causal designs would be beneficial for triangulating these findings.

### Implications

Population-wide initiatives targeting cardiorespiratory fitness early in life might hold promise in contributing to reducing future cancer mortality burden. Such initiatives may be particularly warranted given that population levels of cardiorespiratory fitness are trending downwards across all ages, and especially in young people.^11–13^ While more causal data is needed before definitive policy statements can be made, the influence of behavioural, environmental, and genetic confounders shared between full siblings appear to only partly diminish the public health potential of such interventions.

Taken at face value, the hypothetically preventable share of cancer mortality at 65 years of age, should everyone from the bottom quartile be moved to the second lowest quartile, was estimated to be 16.5% when comparing unrelated individuals in the population and 11.3% when comparing full siblings. This shift assumed that those with the lowest fitness would increase their fitness level by 14% on average. This corresponds roughly to the effect seen in exercise interventions,^55^ although it is unclear to what extent that findings from controlled and typically supervised exercise interventions can be extrapolated to large-scale public health and community interventions.

## Conclusion

This study suggests that high cardiorespiratory fitness in adolescence is associated with lower risk of overall cancer mortality in late adulthood after accounting for behavioural, environmental, and genetic confounders shared by full siblings. Furthermore, our findings provide evidence suggesting that epidemiological studies on this topic may yield biased conclusions if not conducted carefully, as observed excess risks of certain cancers may stem from methodological biases rather than indicating genuine cause for concern.

## Footnotes

### Contributors

MB and VHA conceived the study. MB, MN, PH, and VHA designed the study. PN acquired the ethical permission and the data. MB performed the statistical analyses and drafted the manuscript. VHA verified the underlying data. PN and VHA performed data managing. PN and VHA are joint senior authors. All authors interpreted the data. All authors critically revised the manuscript for intellectual content. The corresponding authors MB and VHA attests that all listed authors meet authorship criteria and that no others meeting the criteria have been omitted. MB and VHA is the manuscript’s guarantors and accept full responsibility for the conduct of the study, had access to the data, and controlled the decision to publish. All authors reviewed and approved the manuscript for submission.

### Ethical approval

This study was approved by the Regional Ethical Review Board in Umeå (no. 2010-113-31M).

### Data sharing

The data in this study are not available to the public and will not be shared according to regulations under Swedish law. Researchers interested in obtaining the data may seek ethical approvals and inquire through Statistics Sweden. For further advice see: https://www.scb.se/en/services/guidance-for-researchers-and-universities/.

### Transparency statement

The lead author MB and the last author VHA (the manuscript’s guarantors) affirm that the manuscript is an honest, accurate, and transparent account of the study being reported; that no important aspects of the study have been omitted; and that any discrepancies from the study as planned (and, if relevant, registered) have been explained.

### Dissemination to participants and related patients and public communities

The findings of this study will be disseminated to the public through a press release, popular science articles and seminars.

### Copyright/licence for publication

The Corresponding Authors have the right to grant on behalf of all authors and do grant on behalf of all authors, a worldwide licence to the Publishers and its licensees in perpetuity, in all forms, formats and media (whether known now or created in the future), to i) publish, reproduce, distribute, display and store the Contribution, ii) translate the Contribution into other languages, create adaptations, reprints, include within collections and create summaries, extracts and/or, abstracts of the Contribution, iii) create any other derivative work(s) based on the Contribution, iv) to exploit all subsidiary rights in the Contribution, v) the inclusion of electronic links from the Contribution to third party material where-ever it may be located; and, vi) licence any third party to do any or all of the above.

## Data Availability

The data in this study are not available to the public and will not be shared ac-cording to regulations under Swedish law. Researchers interested in obtaining the data may seek ethical approvals and inquire through Statistics Sweden. For further advice see: https://www.scb.se/en/services/guidance-for-researchers-and-universities/.

