## Supplementary material for "Adolescent Cardiorespiratory Fitness and Risk of Cancer in Late Adulthood: Nationwide Sibling-Controlled Cohort Study"

Marcel Ballin, associated researcher[1,2,a], Daniel Berglind, associate professor[2,3,4], Pontus Henriksson, associate professor[5], Martin Neovius, professor[6], Anna Nordström, researcher[7,8], Francisco B. Ortega, professor[9,10], Elina Sillanpää, associate professor[10,11], Peter Nordström, professor[1,b], Viktor H. Ahlqvist, postdoctoral researcher[1,12,13,a,b]

[1]Department of Public Health and Caring Sciences, Clinical Geriatrics, Uppsala University, Uppsala, Sweden.

[2]Centre for Epidemiology and Community Medicine, Region Stockholm, Stockholm, Sweden.

[3]Department of Global Public Health, Karolinska Institutet, Stockholm, Sweden.

[4]Center for Wellbeing, Welfare and Happiness, Stockholm School of Economics, Stockholm, Sweden.

[5]Department of Health, Medicine and Caring Sciences, Linköping University, Linköping, Sweden.

[6]Department of Medicine, Clinical Epidemiology Division, Karolinska Institutet, Stockholm, Sweden.

[7]Department of Medical Sciences, Uppsala University, Uppsala, Sweden.

[8]School of Sports Science, UiT The Arctic University of Norway, Tromsø, Norway.

[9]Department of Physical Education and Sports, Faculty of Sport Sciences, Sport and Health University Research Institute (iMUDS), University of Granada; CIBEROBN, ISCIII, Granada, Andalucía, Spain.

[10]Faculty of Sport and Health Sciences, University of Jyväskylä, Jyväskylä, Finland.

[11]Wellbeing Services County of Central Finland, Jyväskylä, Finland.

[12]Department of Biomedicine, Aarhus University, Aarhus, Denmark.

[13]Institute of Environmental Medicine, Karolinska Institutet, Stockholm, Sweden.

[a]Dr. Ballin and Dr. Ahlqvist are joint corresponding authors.

[b]Prof. Nordström and Dr. Ahlqvist are joint senior authors.

**Supplementary table 1. Diagnostic codes used to define the outcomes in the study**

| Cancer outcomes | Code Type* | Codes |
|---|---|---|
| Head and neck | ICD-10 | C00-C14, C30-C32 |
| | ICD-9 | 140-149 |
| | ICD-8 | 140-149 |
| Oesophagus | ICD-10 | C15 |
| | ICD-9 | 150 |
| | ICD-8 | 150 |
| Lung | ICD-10 | C34 |
| | ICD-9 | 162 |
| | ICD-8 | 162 |
| Stomach | ICD-10 | C16 |
| | ICD-9 | 151 |
| | ICD-8 | 151 |
| Pancreas | ICD-10 | C25 |
| | ICD-9 | 157 |
| | ICD-8 | 157 |
| Liver, bile ducts, and gallbladder | ICD-10 | C22-C24 |
| | ICD-9 | 155-156 |
| | ICD-8 | 155-156 |
| Colon | ICD-10 | C18 |
| | ICD-9 | 153 |
| | ICD-8 | 153 |
| Rectum | ICD-10 | C19-C20 |
| | ICD-9 | 154 |
| | ICD-8 | 154 |
| Kidney | ICD-10 | C64 |
| | ICD-9 | 189 |
| | ICD-8 | 189 |
| Prostate | ICD-10 | C61 |
| | ICD-9 | 185 |
| | ICD-8 | 185 |
| Bladder | ICD-10 | C67 |
| | ICD-9 | 188 |
| | ICD-8 | 188 |
| Myeloma | ICD-10 | C90 |
| | ICD-9 | 203 |
| | ICD-8 | 203 |
| Melanoma skin | ICD-10 | C43 |
| | ICD-9 | 172 |
| | ICD-8 | 172 |
| Non-melanoma skin | ICD-10 | C44 |
| | ICD-9 | 173 |
| | ICD-8 | 173 |
| Overall cancer | ICD-10 | C00-C97 |
| | ICD-9 | 140-175, 179-208 |
| | ICD-8 | 140-209 |

ICD = International Classification of Diseases.
*Codes from all versions (8,9,10) were available for the ascertainment of cancer mortality but codes from versions 8 and 9 were not available for the ascertainment of cancer diagnosis as we did not have access to those codes in the National Patient Register. However, we deem the number of such cases to have been very small given that most of the years of follow-up were covered using ICD-10. Moreover, ICD-8 and 9 are only available in inpatient care and are not validated to the same extent as ICD-10.

**Supplementary table 2. Follow-up time, number of events, and numbers censored in cohort and sibling analysis.**

| Cancer outcome | Cohort analysis (N=1 124 049) | Sibling analysis (N=477 453) |
|---|---|---|
| **Any cancer diagnosis** | | |
| Follow-up time, median (range) | 37.7 (0.1-51.4) | 38.3 (0.1-51.4) |
| Events | 98 410 (8.8) | 41 293 (8.7) |
| Death from non-cancer causes | 47 320 (4.2) | 18 579 (3.9) |
| Emigration | 75 786 (6.7) | 31 195 (6.5) |
| End of follow-up | 902 533 (80.3) | 386 386 (80.9) |
| **Any cancer mortality** | | |
| Follow-up time, median (range) | 38.3 (0.1-51.4) | 38.9 (0.1-51.4) |
| Events | 16 789 (1.5) | 6908 (1.5) |
| Death from non-cancer causes | 48 122 (4.3) | 18 912 (4.0) |
| Emigration | 76 274 (6.8) | 31 377 (6.6) |
| End of follow-up | 982 864 (87.4) | 420 256 (88.0) |
| **Head and neck** | | |
| Follow-up time, median (range) | 38.3 (0.1-51.4) | 38.9 (0.1-51.4) |
| Events | 4026 (0.4) | 1692 (0.4) |
| Death from other causes | 63 867 (5.7) | 25 385 (5.3) |
| Emigration | 76 251 (6.8) | 31 370 (6.6) |
| End of follow-up | 979 905 (87.2) | 419 006 (87.8) |
| **Oesophagus** | | |
| Follow-up time, median (range) | 38.3 (0.1-51.4) | 38.9 (0.1-51.4) |
| Events | 1178 (0.1) | 464 (0.1) |
| Death from other causes | 64 069 (5.7) | 25 494 (5.3) |
| Emigration | 76 274 (6.8) | 31 377 (6.6) |
| End of follow-up | 982 528 (87.4) | 420 118 (88.0) |
| **Lung** | | |
| Follow-up time, median (range) | 38.3 (0.1-51.4) | 38.9 (0.1-51.4) |
| Events | 3131 (0.3) | 1263 (0.3) |
| Death from other causes | 62 755 (5.6) | 24 971 (5.2) |
| Emigration | 76 270 (6.8) | 31 375 (6.6) |
| End of follow-up | 981 893 (87.4) | 419 844 (87.9) |
| **Stomach** | | |
| Follow-up time, median (range) | 38.3 (0.1-51.4) | 38.9 (0.1-51.4) |
| Events | 1430 (0.1) | 599 (0.1) |
| Death from other causes | 63 986 (5.7) | 25 443 (5.3) |
| Emigration | 76 272 (6.8) | 31 375 (6.6) |
| End of follow-up | 982 361 (87.4) | 420 036 (88.0) |
| **Pancreas** | | |
| Follow-up time, median (range) | 38.3 (0.1-51.4) | 38.9 (0.1-51.4) |
| Events | 2255 (0.2) | 943 (0.2) |
| Death from other causes | 63 231 (5.6) | 25 134 (5.3) |
| Emigration | 76 273 (6.8) | 31 377 (6.6) |
| End of follow-up | 982 290 (87.4) | 419 999 (88.0) |
| **Liver, bile ducts, and gallbladder** | | |
| Follow-up time, median (range) | 38.3 (0.1-51.4) | 38.9 (0.1-51.4) |
| Events | 2246 (0.2) | 925 (0.2) |
| Death from other causes | 63 322 (5.6) | 25 160 (5.3) |
| Emigration | 76 270 (6.8) | 31 375 (6.6) |
| End of follow-up | 982 211 (87.4) | 419 993 (88.0) |
| **Colon** | | |
| Follow-up time, median (range) | 38.3 (0.1-51.4) | 38.9 (0.1-51.4) |
| Events | 5320 (0.5) | 2177 (0.5) |
| Death from other causes | 63 009 (5.6) | 25 046 (5.3) |
| Emigration | 76 259 (6.8) | 31 374 (6.6) |
| End of follow-up | 979 461 (87.1) | 418 856 (87.7) |
| **Rectum** | | |
| Follow-up time, median (range) | 38.3 (0.1-51.4) | 38.9 (0.1-51.4) |
| Events | 3917 (0.4) | 1625 (0.3) |
| Death from other causes | 63 684 (5.7) | 25 304 (5.3) |
| Emigration | 76 267 (6.8) | 31 372 (6.6) |
| End of follow-up | 980 181 (87.2) | 419 152 (87.8) |
| **Kidney** | | |
| Follow-up time, median (range) | 38.3 (0.1-51.4) | 38.9 (0.1-51.4) |
| Events | 2741 (0.2) | 1151 (0.2) |
| Death from other causes | 64 174 (5.7) | 25 519 (5.3) |
| Emigration | 76 267 (6.8) | 31 373 (6.6) |
| End of follow-up | 980 867 (87.3) | 419 410 (87.8) |
| **Prostate** | | |
| Follow-up time, median (range) | 38.2 (0.1-51.4) | 38.8 (0.1-51.4) |
| Events | 24 225 (2.2) | 10 042 (2.1) |
| Death from other causes | 63 387 (5.6) | 25 229 (5.3) |

| | | |
|---|---|---|
| Emigration | 76 211 (6.8) | 31 353 (6.6) |
| End of follow-up | 960 225 (85.4) | 410 829 (86.1) |
| **Bladder** | | |
| Follow-up time, median (range) | 38.3 (0.1-51.4) | 38.9 (0.1-51.4) |
| Events | 3490 (0.3) | 1432 (0.3) |
| Death from other causes | 64 374 (5.7) | 25 594 (5.4) |
| Emigration | 76 256 (6.8) | 31 367 (6.6) |
| End of follow-up | 979 929 (87.2) | 419 060 (87.8) |
| **Myeloma** | | |
| Follow-up time, median (range) | 38.3 (0.1-51.4) | 38.9 (0.1-51.4) |
| Events | 1460 (0.1) | 580 (0.1) |
| Death from other causes | 64 446 (5.7) | 25 643 (5.4) |
| Emigration | 76 271 (6.8) | 31 374 (6.6) |
| End of follow-up | 981 872 (87.4) | 419 856 (87.9) |
| **Melanoma** | | |
| Follow-up time, median (range) | 38.2 (0.1-51.4) | 38.8 (0.1-51.4) |
| Events | 10 026 (0.9) | 4216 (0.9) |
| Death from other causes | 63 730 (5.7) | 25 360 (5.3) |
| Emigration | 76 207 (6.8) | 31 359 (6.6) |
| End of follow-up | 974 086 (86.7) | 416 518 (87.2) |
| **Non-melanoma** | | |
| Follow-up time, median (range) | 38.1 (0.1-51.4) | 38.7 (0.1-51.4) |
| Events | 27 302 (2.4) | 11 509 (2.4) |
| Death from other causes | 63 579 (5.7) | 25 284 (5.3) |
| Emigration | 76 111 (6.8) | 31 313 (6.6) |
| End of follow-up | 957 057 (85.1) | 409 347 (85.7) |

Number of events and numbers censored are shown as n (%).

**Supplementary table 3. Hazard ratios for cancer by quartiles of cardiorespiratory fitness in cohort and sibling analysis.**

| | Cohort analysis (N=1 124 049) | | Sibling analysis (N=477 453) | |
|---|---|---|---|---|
| **Cancer outcome by quartiles of cardiorespiratory fitness** | **Cases, n (%)** | **HR (95% CI)** | **Cases, n (%)** | **HR (95% CI)** |
| **Overall cancer diagnosis** | | | | |
| Q1 | 33 673 (12.0) | Ref. | 13 621 (11.5) | Ref. |
| Q2 | 28 078 (9.8) | 1.01 (0.99, 1.02) | 11 693 (9.6) | 1.00 (0.96, 1.03) |
| Q3 | 20 523 (7.4) | 1.03 (1.01, 1.05) | 8990 (7.6) | 1.02 (0.97, 1.06) |
| Q4 | 16 136 (5.8) | 1.08 (1.06, 1.11) | 6989 (5.9) | 1.00 (0.95, 1.06) |
| **Overall cancer mortality** | | | | |
| Q1 | 7093 (2.5) | Ref. | 2790 (2.4) | Ref. |
| Q2 | 4810 (1.7) | 0.83 (0.80, 0.86) | 1981 (1.6) | 0.88 (0.81, 0.96) |
| Q3 | 2960 (1.1) | 0.76 (0.73, 0.80) | 1300 (1.1) | 0.85 (0.77, 0.95) |
| Q4 | 1926 (0.7) | 0.71 (0.67, 0.76) | 837 (0.7) | 0.78 (0.68, 0.89) |
| **Site-specific cancers** | | | | |
| Head and neck | | | | |
| Q1 | 1539 (0.6) | Ref. | 629 (0.5) | Ref. |
| Q2 | 1148 (0.4) | 0.85 (0.79, 0.93) | 474 (0.4) | 0.94 (0.78, 1.12) |
| Q3 | 782 (0.3) | 0.80 (0.73, 0.88) | 350 (0.3) | 0.84 (0.68, 1.04) |
| Q4 | 557 (0.2) | 0.75 (0.67, 0.85) | 237 (0.2) | 0.80 (0.62, 1.04) |
| Oesophagus | | | | |
| Q1 | 536 (0.2) | Ref. | 213 (0.2) | Ref. |
| Q2 | 346 (0.1) | 0.75 (0.65, 0.86) | 139 (0.1) | 0.95 (0.69, 1.32) |
| Q3 | 185 (0.07) | 0.60 (0.50, 0.72) | 68 (0.06) | 0.68 (0.45, 1.03) |
| Q4 | 111 (0.04) | 0.53 (0.42, 0.66) | 44 (0.04) | 0.61 (0.36, 1.02) |
| Lung | | | | |
| Q1 | 1562 (0.6) | Ref. | 620 (0.5) | Ref. |
| Q2 | 871 (0.3) | 0.74 (0.68, 0.80) | 350 (0.3) | 0.81 (0.67, 0.98) |
| Q3 | 460 (0.2) | 0.62 (0.55, 0.69) | 188 (0.2) | 0.73 (0.56, 0.93) |
| Q4 | 238 (0.09) | 0.49 (0.42, 0.57) | 105 (0.09) | 0.50 (0.36, 0.70) |
| Stomach | | | | |
| Q1 | 597 (0.2) | Ref. | 234 (0.2) | Ref. |
| Q2 | 405 (0.1) | 0.79 (0.70, 0.90) | 164 (0.1) | 0.90 (0.67, 1.21) |
| Q3 | 256 (0.09) | 0.74 (0.63, 0.86) | 101 (0.08) | 0.69 (0.49, 0.98) |
| Q4 | 172 (0.06) | 0.71 (0.58, 0.86) | 79 (0.07) | 0.90 (0.60, 1.37) |
| Pancreas | | | | |
| Q1 | 925 (0.3) | Ref. | 369 (0.3) | Ref. |
| Q2 | 663 (0.2) | 0.86 (0.78, 0.95) | 270 (0.2) | 1.05 (0.83, 1.33) |
| Q3 | 424 (0.2) | 0.84 (0.74, 0.95) | 191 (0.2) | 0.98 (0.74, 1.30) |
| Q4 | 243 (0.09) | 0.73 (0.62, 0.85) | 113 (0.1) | 0.83 (0.59, 1.18) |
| Liver, bile ducts, and gallbladder | | | | |
| Q1 | 1038 (0.4) | Ref. | 380 (0.3) | Ref. |
| Q2 | 657 (0.2) | 0.77 (0.69, 0.85) | 294 (0.2) | 1.10 (0.86, 1.39) |
| Q3 | 333 (0.1) | 0.60 (0.52, 0.68) | 163 (0.1) | 0.97 (0.73, 1.28) |
| Q4 | 218 (0.08) | 0.59 (0.50, 0.69) | 88 (0.07) | 0.78 (0.54, 1.13) |
| Colon | | | | |
| Q1 | 2001 (0.7) | Ref. | 801 (0.7) | Ref. |
| Q2 | 1543 (0.5) | 0.91 (0.85, 0.97) | 618 (0.5) | 0.89 (0.76, 1.04) |
| Q3 | 1050 (0.4) | 0.84 (0.78, 0.91) | 446 (0.4) | 0.92 (0.76, 1.11) |
| Q4 | 726 (0.3) | 0.75 (0.68, 0.83) | 312 (0.3) | 0.78 (0.62, 0.99) |
| Rectum | | | | |
| Q1 | 1474 (0.5) | Ref. | 615 (0.5) | Ref. |
| Q2 | 1154 (0.4) | 0.94 (0.87, 1.02) | 470 (0.4) | 0.90 (0.75, 1.07) |
| Q3 | 764 (0.3) | 0.88 (0.80, 0.96) | 325 (0.3) | 0.84 (0.68, 1.04) |
| Q4 | 525 (0.2) | 0.81 (0.72, 0.91) | 215 (0.2) | 0.69 (0.53, 0.90) |
| Kidney | | | | |
| Q1 | 1035 (0.4) | Ref. | 416 (0.4) | Ref. |
| Q2 | 801 (0.3) | 0.84 (0.76, 0.92) | 352 (0.3) | 1.01 (0.81, 1.26) |
| Q3 | 529 (0.2) | 0.73 (0.65, 0.82) | 226 (0.2) | 0.91 (0.70, 1.18) |
| Q4 | 376 (0.1) | 0.65 (0.57, 0.75) | 157 (0.1) | 0.86 (0.63, 1.18) |
| Prostate | | | | |
| Q1 | 9678 (3.4) | Ref. | 3887 (3.3) | Ref. |
| Q2 | 7507 (2.6) | 1.04 (1.01, 1.07) | 3061 (2.5) | 1.00 (0.94, 1.08) |
| Q3 | 4449 (1.6) | 1.05 (1.01, 1.09) | 1947 (1.6) | 1.03 (0.94, 1.12) |
| Q4 | 2591 (0.9) | 1.10 (1.05, 1.16) | 1147 (1.0) | 1.01 (0.90, 1.13) |
| Bladder | | | | |
| Q1 | 1427 (0.5) | Ref. | 589 (0.5) | Ref. |
| Q2 | 1018 (0.4) | 0.90 (0.82, 0.97) | 405 (0.3) | 0.98 (0.81, 1.19) |
| Q3 | 667 (0.2) | 0.89 (0.81, 0.99) | 270 (0.2) | 0.85 (0.68, 1.05) |
| Q4 | 378 (0.1) | 0.75 (0.65, 0.85) | 168 (0.1) | 0.89 (0.66, 1.19) |
| Myeloma | | | | |
| Q1 | 516 (0.2) | Ref. | 199 (0.2) | Ref. |
| Q2 | 429 (0.2) | 0.99 (0.86, 1.13) | 166 (0.1) | 0.76 (0.55, 1.04) |

| | | | | |
|---|---|---|---|---|
| Q3 | 301 (0.1) | 1.00 (0.86, 1.17) | 123 (0.1) | 0.83 (0.57, 1.20) |
| Q4 | 214 (0.08) | 1.00 (0.83, 1.20) | 92 (0.08) | 0.81 (0.50, 1.29) |
| Melanoma skin | | | | |
| Q1 | 2622 (0.9) | Ref. | 1031 (0.9) | Ref. |
| Q2 | 2682 (0.9) | 1.13 (1.07, 1.19) | 1113 (0.9) | 1.08 (0.96, 1.22) |
| Q3 | 2396 (0.9) | 1.28 (1.21, 1.37) | 1067 (0.9) | 1.22 (1.06, 1.40) |
| Q4 | 2326 (0.8) | 1.50 (1.41, 1.61) | 1005 (0.9) | 1.30 (1.11, 1.52) |
| Non-melanoma skin | | | | |
| Q1 | 8441 (3.0) | Ref. | 3374 (2.9) | Ref. |
| Q2 | 7726 (2.7) | 1.13 (1.09, 1.17) | 3192 (2.6) | 1.03 (0.96, 1.11) |
| Q3 | 5957 (2.2) | 1.22 (1.18, 1.27) | 2689 (2.3) | 1.09 (1.01, 1.19) |
| Q4 | 5178 (1.9) | 1.44 (1.37, 1.50) | 2254 (1.9) | 1.09 (0.99, 1.20) |

CI = confidence interval. HR = hazard ratio. Q = quartile. HRs are adjusted for age at conscription, year of conscription, body mass index, parental education, and parental income. In both cohorts, the median (range) of $W_{max}$ in Q1 was 217 (100-236), in Q2 it was 253 (237-270), in Q3 it was 290 (271-312), in Q4 it was 339 (313-999).

**Supplementary table 4. Standardised cumulative incidence of cancer at 65 years of age by quartiles of cardiorespiratory fitness in cohort and sibling analysis.**

| | Cohort analysis (N=1 124 049) | | Sibling analysis (N=477 453) | |
|---|---|---|---|---|
| **Cancer outcome by quartiles of cardiorespiratory fitness** | **Standardised incidence at age 65 y, % (95% CI)** | **Difference, pp (95% CI)** | **Standardised incidence at age 65 y, % (95% CI)** | **Difference, pp (95% CI)** |
| **Overall cancer diagnosis** | | | | |
| Q1 | 18.1 (17.8, 18.3) | Ref. | 17.8 (17.3, 18.4) | Ref. |
| Q2 | 18.6 (18.4, 18.9) | 0.6 (0.2, 0.9) | 17.8 (17.3, 18.3) | -0.004 (-0.9, 0.8) |
| Q3 | 19.0 (18.6, 19.2) | 0.9 (0.5, 1.3) | 18.1 (17.5, 18.6) | 0.2 (-0.7, 1.2) |
| Q4 | 19.5 (19.2, 19.9) | 1.5 (1.0, 1.9) | 17.9 (17.3, 18.6) | 0.08 (-0.9, 1.1) |
| **Overall cancer mortality** | | | | |
| Q1 | 3.0 (2.9, 3.1) | Ref. | 2.8 (2.6, 3.0) | Ref. |
| Q2 | 2.2 (2.1, 2.3) | -0.8 (-0.9, -0.7) | 2.3 (2.1, 2.4) | -0.5 (-0.8, -0.2) |
| Q3 | 2.0 (2.0, 2.1) | -1.0 (-1.1, -0.8) | 2.2 (2.0, 2.4) | -0.6 (-0.9, -0.3) |
| Q4 | 1.9 (1.8, 2.0) | -1.1 (-1.2, -0.9) | 2.1 (1.8, 2.3) | -0.7 (-1.1, -0.4) |
| **Site-specific cancers** | | | | |
| Head and neck | | | | |
| Q1 | 0.8 (0.7, 0.8) | Ref. | 0.7 (0.6, 0.8) | Ref. |
| Q2 | 0.6 (0.6, 0.6) | -0.2 (-0.3, -0.1) | 0.6 (0.5, 0.7) | -0.1 (-0.3, 0.06) |
| Q3 | 0.6 (0.5, 0.6) | -0.2 (-0.3, -0.1) | 0.6 (0.5, 0.7) | -0.2 (-0.3, -0.02) |
| Q4 | 0.5 (0.5, 0.6) | -0.2 (-0.3, -0.2) | 0.5 (0.4, 0.7) | -0.2 (-0.4, -0.02) |
| Oesophagus | | | | |
| Q1 | 0.3 (0.2, 0.3) | Ref. | 0.2 (0.2, 0.3) | Ref. |
| Q2 | 0.2 (0.1, 0.2) | -0.1 (-0.2, -0.08) | 0.2 (0.1, 0.2) | -0.05 (-0.1, 0.04) |
| Q3 | 0.1 (0.1, 0.2) | -0.1 (-0.2, -0.09) | 0.1 (0.1, 0.2) | -0.08 (-0.2, 0.001) |
| Q4 | 0.1 (0.1, 0.2) | -0.2 (-0.2, -0.1) | 0.1 (0.1, 0.2) | -0.09 (-0.2, -0.004) |
| Lung | | | | |
| Q1 | 0.6 (0.6, 0.7) | Ref. | 0.6 (0.5, 0.7) | Ref. |
| Q2 | 0.4 (0.3, 0.4) | -0.3 (-0.3, -0.2) | 0.4 (0.3, 0.5) | -0.2 (-0.3, -0.07) |
| Q3 | 0.3 (0.3, 0.4) | -0.3 (-0.4, -0.3) | 0.4 (0.3, 0.4) | -0.2 (-0.4, -0.09) |
| Q4 | 0.3 (0.2, 0.3) | -0.4 (-0.4, -0.3) | 0.3 (0.2, 0.3) | -0.3 (-0.5, -0.2) |
| Stomach | | | | |
| Q1 | 0.3 (0.2, 0.3) | Ref. | 0.3 (0.2, 0.3) | Ref. |
| Q2 | 0.2 (0.2, 0.2) | -0.08 (-0.1, -0.05) | 0.2 (0.2, 0.3) | -0.05 (-0.1, 0.04) |
| Q3 | 0.2 (0.2, 0.2) | -0.09 (-0.1, -0.05) | 0.2 (0.1, 0.2) | -0.09 (-0.2, 0.002) |
| Q4 | 0.2 (0.1, 0.2) | -0.1 (-0.1, -0.06) | 0.2 (0.2, 0.3) | -0.05 (0.2, 0.06) |
| Pancreas | | | | |
| Q1 | 0.5 (0.4, 0.5) | Ref. | 0.4 (0.3, 0.5) | Ref. |
| Q2 | 0.4 (0.3, 0.4) | -0.1 (-0.2, -0.05) | 0.4 (0.3, 0.5) | 0.0002 (-0.1, 0.1) |
| Q3 | 0.4 (0.3, 0.4) | -0.1 (-0.2, -0.05) | 0.4 (0.3, 0.5) | -0.02 (-0.2, 0.1) |
| Q4 | 0.3 (0.3, 0.4) | -0.2 (-0.2, -0.08) | 0.4 (0.3, 0.5) | -0.07 (-0.2, 0.09) |
| Liver, bile ducts, and gallbladder | | | | |
| Q1 | 0.5 (0.5, 0.6) | Ref. | 0.4 (0.3, 0.5) | Ref. |
| Q2 | 0.3 (0.3, 0.3) | -0.2 (-0.3, -0.1) | 0.4 (0.3, 0.5) | 0.01 (-0.1, 0.1) |
| Q3 | 0.3 (0.2, 0.3) | -0.3 (-0.3, -0.2) | 0.4 (0.3, 0.4) | -0.02 (-0.2, 0.1) |
| Q4 | 0.3 (0.2, 0.3) | -0.3 (-0.3, -0.2) | 0.3 (0.2, 0.4) | -0.08 (-0.2, 0.06) |
| Colon | | | | |
| Q1 | 1.2 (1.1, 1.2) | Ref. | 1.1 (1.0, 1.3) | Ref. |
| Q2 | 0.9 (0.9, 1.0) | -0.2 (-0.3, -0.1) | 0.9 (0.8, 1.0) | -0.2 (-0.4, -0.02) |
| Q3 | 0.9 (0.8, 1.0) | -0.3 (-0.4, -0.2) | 0.9 (0.8, 1.1) | -0.2 (-0.4, 0.07) |
| Q4 | 0.8 (0.8, 0.9) | -0.3 (-0.5, -0.2) | 0.8 (0.7, 1.0) | -0.3 (-0.5, -0.02) |
| Rectum | | | | |
| Q1 | 0.9 (0.8, 0.9) | Ref. | 0.9 (0.7, 1.0) | Ref. |
| Q2 | 0.7 (0.7, 0.8) | -0.1 (-0.2, -0.04) | 0.7 (0.6, 0.8) | -0.2 (-0.4, 0.003) |
| Q3 | 0.7 (0.6, 0.8) | -0.2 (-0.3, -0.07) | 0.6 (0.5, 0.8) | -0.2 (-0.4, 0.01) |
| Q4 | 0.7 (0.6, 0.7) | -0.2 (-0.3, -0.09) | 0.6 (0.5, 0.7) | -0.3 (-0.5, -0.09) |
| Kidney | | | | |
| Q1 | 0.6 (0.6, 0.7) | Ref. | 0.5 (0.4, 0.6) | Ref. |
| Q2 | 0.4 (0.4, 0.5) | -0.2 (-0.3, -0.1) | 0.5 (0.4, 0.6) | -0.03 (-0.2, 0.1) |
| Q3 | 0.4 (0.4, 0.5) | -0.2 (-0.3, -0.2) | 0.5 (0.4, 0.6) | -0.06 (-0.2, 0.1) |
| Q4 | 0.4 (0.3, 0.4) | -0.3 (-0.4, -0.2) | 0.4 (0.3, 0.6) | -0.08 (-0.3, 0.09) |
| Prostate | | | | |
| Q1 | 4.9 (4.7, 5.1) | Ref. | 5.0 (4.7, 5.4) | Ref. |
| Q2 | 5.3 (5.1, 5.5) | 0.4 (0.2, 0.6) | 5.1 (4.8, 5.5) | 0.06 (-0.4, 0.6) |
| Q3 | 5.4 (5.1, 5.6) | 0.4 (0.2, 0.7) | 5.2 (4.8, 5.6) | 0.2 (-0.4, 0.7) |
| Q4 | 5.5 (5.3, 5.8) | 0.6 (0.3, 0.9) | 5.1 (4.7, 5.6) | 0.08 (-0.6, 0.8) |
| Bladder | | | | |
| Q1 | 0.7 (0.6, 0.7) | Ref. | 0.6 (0.5, 0.7) | Ref. |
| Q2 | 0.5 (0.5, 0.6) | -0.1 (-0.2, -0.06) | 0.5 (0.5, 0.6) | -0.05 (-0.2, 0.09) |
| Q3 | 0.5 (0.5, 0.6) | -0.1 (-0.2, -0.05) | 0.5 (0.4, 0.6) | -0.1 (-0.3, 0.04) |
| Q4 | 0.5 (0.4, 0.5) | -0.2 (-0.3, -0.1) | 0.5 (0.4, 0.6) | -0.09 (-0.3, 0.09) |
| Myeloma | | | | |
| Q1 | 0.3 (0.2, 0.3) | Ref. | 0.3 (0.2, 0.4) | Ref. |

| | | | | |
|---|---|---|---|---|
| Q2 | 0.3 (0.2, 0.3) | -0.004 (-0.05, 0.04) | 0.2 (0.2, 0.3) | -0.09 (-0.2, 0.02) |
| Q3 | 0.3 (0.2, 0.3) | 0.00001 (-0.05, 0.05) | 0.2 (0.2, 0.3) | -0.08 (-0.2, 0.05) |
| Q4 | 0.3 (0.2, 0.3) | 0.001 (-0.06, 0.06) | 0.2 (0.2, 0.3) | -0.08 (-0.2, 0.06) |
| Melanoma skin | | | | |
| Q1 | 1.7 (1.5, 1.7) | Ref. | 1.7 (1.5, 1.8) | Ref. |
| Q2 | 2.2 (2.1, 2.3) | 0.6 (0.5, 0.8) | 2.1 (1.8, 2.3) | 0.4 (0.06, 0.7) |
| Q3 | 2.4 (2.3, 2.6) | 0.8 (0.6, 1.0) | 2.2 (2.0, 2.5) | 0.6 (0.2, 0.9) |
| Q4 | 2.7 (2.5, 2.8) | 1.0 (0.9, 1.3) | 2.3 (2.0, 2.6) | 0.6 (0.2, 1.1) |
| Non-melanoma skin | | | | |
| Q1 | 4.4 (4.2, 4.5) | Ref. | 4.7 (4.4, 5.0) | Ref. |
| Q2 | 5.7 (5.6, 5.9) | 1.4 (1.1, 1.6) | 5.1 (4.8, 5.4) | 0.4 (-0.1, 0.9) |
| Q3 | 6.1 (5.9, 6.3) | 1.7 (1.4, 2.0) | 5.3 (5.0, 5.7) | 0.6 (0.03, 0.1) |
| Q4 | 6.7 (6.5, 7.0) | 2.4 (2.1, 2.7) | 5.3 (4.9, 5.7) | 0.6 (-0.05, 0.1) |

CI = confidence interval. Q = quartile. All estimates are adjusted for age at conscription, year of conscription, body mass index, parental education, and parental income. In both cohorts, the median (range) of $W_{max}$ in Q1 was 217 (100-236), in Q2 it was 253 (237-270), in Q3 it was 290 (271-312), in Q4 it was 339 (313-999).

**Supplementary table 5. Population-attributable fraction for cancer at 65 years of age by cardiorespiratory fitness in cohort and sibling analysis, when considering a moderate (shifting those in quartile 1 to quartile 2) or a major intervention (shifting everyone to quartile 4).**

| | Moderate intervention | | Major intervention | |
|---|---|---|---|---|
| | **Cohort analysis (N=1 124 049)** | **Sibling analysis (N=477 453)** | **Cohort analysis (N=1 124 049)** | **Sibling analysis (N=477 453)** |
| **Cancer outcome** | **PAF, % (95% CI)** | **PAF, % (95% CI)** | **PAF, % (95% CI)** | **PAF, % (95% CI)** |
| **Overall cancer diagnosis** | -2.0 (-3.0, -1.0) | -0.1 (-2.5, 2.2) | -5.3 (-6.8, -3.7) | -0.2 (-3.6, 3.6) |
| **Overall cancer mortality** | 16.5 (14.4, 18.6) | 11.3 (6.1, 16.6) | 22.6 (19.2, 26.1) | 17.2 (8.8, 25.5) |
| **Site specific cancers** | | | | |
| Head and neck | 14.0 (9.5, 18.4) | 9.3 (-1.9, 20.6) | 18.9 (12.0, 25.8) | 15.5 (-1.2, 32.2) |
| Oesophagus | 26.4 (19.8, 33.1) | 16.2 (-2.5, 34.9) | 38.0 (27.0, 49.0) | 32.0 (5.3, 58.7) |
| Lung | 27.9 (23.8, 32.0) | 22.9 (12.7, 33.1) | 42.8 (35.7, 50.0) | 41.4 (26.5, 56.2) |
| Stomach | 18.6 (11.6, 25.6) | 11.6 (-5.9, 29.1) | 23.3 (11.8, 34.8) | 7.1 (-22.2, 36.3) |
| Pancreas | 13.6 (7.7, 19.5) | 1.6 (-14.7, 17.9) | 21.4 (11.7, 31.0) | 12.9 (-9.5, 35.3) |
| Liver, bile ducts, and gallbladder | 24.8 (20.0, 29.8) | 1.7 (-14.6, 17.9) | 32.9 (24.3, 41.4) | 17.9 (-5.8, 41.5) |
| Colon | 11.1 (7.1, 15.1) | 9.7 (0.2, 19.3) | 18.7 (12.8, 24.6) | 16.4 (2.5, 30.3) |
| Rectum | 8.2 (3.4, 13.1) | 12.7 (2.2, 23.2) | 14.5 (7.2, 21.7) | 26.7 (9.4, 37.9) |
| Kidney | 17.6 (12.6, 22.7) | 3.9 (-10.6, 18.3) | 26.5 (19.2, 33.8) | 10.6 (-10.3, 31.5) |
| Prostate | -4.2 (-6.4, -1.9) | -0.8 (-5.9, 4.3) | -7.2 (-10.1, -3.4) | -0.7 (-9.9, 7.4) |
| Bladder | 10.9 (5.9, 15.8) | 6.0 (-6.6, 18.5) | 20.1 (12.1, 28.1) | 9.1 (-11.7, 29.8) |
| Myeloma | 0.5 (-8.5, 9.4) | 16.0 (-2.3, 34.3) | 0.2 (-13.5, 13.9) | 14.2 (-13.9, 42.3) |
| Melanoma skin | -18.2 (-22.7, -13.7) | -12.9 (-21.6, -2.2) | -32.3 (-38.5, -26.2) | -19.1 (-32.2, -6.0) |
| Non-melanoma skin | -16.0 (-18.5, -13.5) | -4.3 (-9.5, 0.9) | -28.8 (-32.5, -25.0) | -5.9 (-13.1, 1.3) |

CI = confidence interval. PAF = population-attributable fraction. All estimates were adjusted for age at conscription, year of conscription, body mass index, parental education, and parental income. Negative estimates imply a theoretically increased risk of the outcome in the population should the relevant intervention be carried out

**Supplementary table 6. Hazard ratios for cancer by quartiles of cardiorespiratory fitness in cohort analysis (as reported in the main article), in the sibling cohort using standard analysis, and in the sibling cohort using sibling analysis (as reported in the main article).**

| Cancer outcome by quartiles of cardiorespiratory fitness | Total cohort (standard) analysis as reported in main article (N=1 124 049) HR (95% CI) | Standard analysis replicated in sibling cohort (N=477 453) HR (95% CI) | Sibling analysis as reported in main article (N=477 453) HR (95% CI) |
|---|---|---|---|
| **Overall cancer diagnosis** | | | |
| Q1 | Ref. | Ref. | Ref. |
| Q2 | 1.01 (0.99, 1.02) | 1.00 (0.98, 1.03) | 1.00 (0.96, 1.03) |
| Q3 | 1.03 (1.01, 1.05) | 1.06 (1.03, 1.09) | 1.02 (0.97, 1.06) |
| Q4 | 1.08 (1.06, 1.11) | 1.09 (1.06, 1.13) | 1.00 (0.95, 1.06) |
| **Overall cancer mortality** | | | |
| Q1 | Ref. | Ref. | Ref. |
| Q2 | 0.83 (0.80, 0.86) | 0.83 (0.78, 0.88) | 0.88 (0.81, 0.96) |
| Q3 | 0.76 (0.73, 0.80) | 0.77 (0.72, 0.82) | 0.85 (0.77, 0.95) |
| Q4 | 0.71 (0.67, 0.76) | 0.68 (0.63, 0.74) | 0.78 (0.68, 0.89) |
| **Site-specific cancers** | | | |
| Head and neck | | | |
| Q1 | Ref. | Ref. | Ref. |
| Q2 | 0.85 (0.79, 0.93) | 0.83 (0.73, 0.93) | 0.94 (0.78, 1.12) |
| Q3 | 0.80 (0.73, 0.88) | 0.80 (0.70, 0.92) | 0.84 (0.68, 1.04) |
| Q4 | 0.75 (0.67, 0.85) | 0.69 (0.58, 0.81) | 0.80 (0.62, 1.04) |
| Oesophagus | | | |
| Q1 | Ref. | Ref. | Ref. |
| Q2 | 0.75 (0.65, 0.86) | 0.73 (0.58, 0.91) | 0.95 (0.69, 1.32) |
| Q3 | 0.60 (0.50, 0.72) | 0.52 (0.39, 0.69) | 0.68 (0.45, 1.03) |
| Q4 | 0.53 (0.42, 0.66) | 0.48 (0.34, 0.69) | 0.61 (0.36, 1.02) |
| Lung | | | |
| Q1 | Ref. | Ref. | Ref. |
| Q2 | 0.74 (0.68, 0.80) | 0.71 (0.62, 0.81) | 0.81 (0.67, 0.98) |
| Q3 | 0.62 (0.55, 0.69) | 0.57 (0.48, 0.68) | 0.73 (0.56, 0.93) |
| Q4 | 0.49 (0.42, 0.57) | 0.47 (0.37, 0.59) | 0.50 (0.36, 0.70) |
| Stomach | | | |
| Q1 | Ref. | Ref. | Ref. |
| Q2 | 0.79 (0.70, 0.90) | 0.76 (0.62, 0.93) | 0.90 (0.67, 1.21) |
| Q3 | 0.74 (0.63, 0.86) | 0.63 (0.49, 0.80) | 0.69 (0.49, 0.98) |
| Q4 | 0.71 (0.58, 0.86) | 0.67 (0.50, 0.90) | 0.90 (0.60, 1.37) |
| Pancreas | | | |
| Q1 | Ref. | Ref. | Ref. |
| Q2 | 0.86 (0.78, 0.95) | 0.84 (0.71, 0.99) | 1.05 (0.83, 1.33) |
| Q3 | 0.84 (0.74, 0.95) | 0.86 (0.71, 1.04) | 0.98 (0.74, 1.30) |
| Q4 | 0.73 (0.62, 0.85) | 0.74 (0.58, 0.93) | 0.83 (0.59, 1.18) |
| Liver, bile ducts, and gallbladder | | | |
| Q1 | Ref. | Ref. | Ref. |
| Q2 | 0.77 (0.69, 0.85) | 0.87 (0.74, 1.01) | 1.10 (0.86, 1.39) |
| Q3 | 0.60 (0.52, 0.68) | 0.69 (0.57, 0.84) | 0.97 (0.73, 1.28) |
| Q4 | 0.59 (0.50, 0.69) | 0.53 (0.41, 0.69) | 0.78 (0.54, 1.13) |
| Colon | | | |
| Q1 | Ref. | Ref. | Ref. |
| Q2 | 0.91 (0.85, 0.97) | 0.88 (0.79, 0.98) | 0.89 (0.76, 1.04) |
| Q3 | 0.84 (0.78, 0.91) | 0.83 (0.74, 0.95) | 0.92 (0.76, 1.11) |
| Q4 | 0.75 (0.68, 0.83) | 0.74 (0.64, 0.86) | 0.78 (0.62, 0.99) |
| Rectum | | | |
| Q1 | Ref. | Ref. | Ref. |
| Q2 | 0.94 (0.87, 1.02) | 0.88 (0.77, 0.99) | 0.90 (0.75, 1.07) |
| Q3 | 0.88 (0.80, 0.96) | 0.83 (0.71, 0.95) | 0.84 (0.68, 1.04) |
| Q4 | 0.81 (0.72, 0.91) | 0.72 (0.60, 0.86) | 0.69 (0.53, 0.90) |
| Kidney | | | |
| Q1 | Ref. | Ref. | Ref. |
| Q2 | 0.84 (0.76, 0.92) | 0.91 (0.79 1.05) | 1.01 (0.81, 1.26) |
| Q3 | 0.73 (0.65, 0.82) | 0.75 (0.63, 0.90) | 0.91 (0.70, 1.18) |
| Q4 | 0.65 (0.57, 0.75) | 0.65 (0.53, 0.80) | 0.86 (0.63, 1.18) |
| Prostate | | | |
| Q1 | Ref. | Ref. | Ref. |
| Q2 | 1.04 (1.01, 1.07) | 1.01 (0.96, 1.06) | 1.00 (0.94, 1.08) |
| Q3 | 1.05 (1.01, 1.09) | 1.05 (0.99, 1.12) | 1.03 (0.94, 1.12) |
| Q4 | 1.10 (1.05, 1.16) | 1.07 (0.99, 1.15) | 1.01 (0.90, 1.13) |
| Bladder | | | |
| Q1 | Ref. | Ref. | Ref. |
| Q2 | 0.90 (0.82, 0.97) | 0.82 (0.72, 0.93) | 0.98 (0.81, 1.19) |
| Q3 | 0.89 (0.81, 0.99) | 0.80 (0.68, 0.93) | 0.85 (0.68, 1.05) |
| Q4 | 0.75 (0.65, 0.85) | 0.72 (0.59, 0.87) | 0.89 (0.66, 1.19) |
| Myeloma | | | |

| | | | |
|---|---|---|---|
| Q1 | Ref. | Ref. | Ref. |
| Q2 | 0.99 (0.86, 1.13) | 0.97 (0.78, 1.19) | 0.76 (0.55, 1.04) |
| Q3 | 1.00 (0.86, 1.17) | 1.02 (0.80, 1.29) | 0.83 (0.57, 1.20) |
| Q4 | 1.00 (0.83, 1.20) | 1.06 (0.80, 1.41) | 0.81 (0.50, 1.29) |
| Melanoma skin | | | |
| Q1 | Ref. | Ref. | Ref. |
| Q2 | 1.13 (1.07, 1.19) | 1.18 (1.08, 1.29) | 1.08 (0.96, 1.22) |
| Q3 | 1.28 (1.21, 1.37) | 1.43 (1.31, 1.57) | 1.22 (1.06, 1.40) |
| Q4 | 1.50 (1.41, 1.61) | 1.66 (1.50, 1.84) | 1.30 (1.11, 1.52) |
| Non-melanoma skin | | | |
| Q1 | Ref. | Ref. | Ref. |
| Q2 | 1.13 (1.09, 1.17) | 1.15 (1.09, 1.20) | 1.03 (0.96, 1.11) |
| Q3 | 1.22 (1.18, 1.27) | 1.34 (1.26, 1.41) | 1.09 (1.01, 1.19) |
| Q4 | 1.44 (1.37, 1.50) | 1.52 (1.43, 1.62) | 1.09 (0.99, 1.20) |

CI = confidence interval. HR = hazard ratio. Q = quartile. All estimates were adjusted for birth cohort, year of conscription, body mass index, parental education, and parental income. In both cohorts, the median (range) of $W_{max}$ in Q1 was 217 (100-236), in Q2 it was 253 (237-270), in Q3 it was 290 (271-312), in Q4 it was 339 (313-999).

**Supplementary table 7. Hazard ratios for cancer by quartiles of cardiorespiratory fitness in cohort and sibling analysis, with and without adjusting for body mass index.**

| | Cohort analysis (N=1 124 049) | | Sibling analysis (N=477 453) | |
|---|---|---|---|---|
| **Cancer outcome by quartiles of cardiorespiratory fitness** | **HR (95% CI) without adjustment for BMI** | **HR (95% CI) with adjustment for BMI** | **HR (95% CI) without adjustment for BMI** | **HR (95% CI) with adjustment for BMI** |
| **Overall cancer diagnosis** | | | | |
| Q1 | Ref. | Ref. | Ref. | Ref. |
| Q2 | 1.01 (1.00, 1.03) | 1.01 (0.99, 1.02) | 1.00 (0.96, 1.04) | 1.00 (0.96, 1.03) |
| Q3 | 1.04 (1.02, 1.06) | 1.03 (1.01, 1.05) | 1.02 (0.98, 1.06) | 1.02 (0.97, 1.06) |
| Q4 | 1.10 (1.08, 1.12) | 1.08 (1.06, 1.11) | 1.01 (0.96, 1.06) | 1.00 (0.95, 1.06) |
| **Overall cancer mortality** | | | | |
| Q1 | Ref. | Ref. | Ref. | Ref. |
| Q2 | 0.87 (0.84, 0.90) | 0.83 (0.80, 0.86) | 0.90 (0.83, 0.98) | 0.88 (0.81, 0.96) |
| Q3 | 0.81 (0.78, 0.85) | 0.76 (0.73, 0.80) | 0.88 (0.79, 0.97) | 0.85 (0.77, 0.95) |
| Q4 | 0.78 (0.74, 0.82) | 0.71 (0.67, 0.76) | 0.82 (0.72, 0.93) | 0.78 (0.68, 0.89) |
| **Site-specific cancer** | | | | |
| Head and neck | | | | |
| Q1 | Ref. | Ref. | Ref. | Ref. |
| Q2 | 0.90 (0.83, 0.97) | 0.85 (0.79, 0.93) | 0.93 (0.78, 1.11) | 0.94 (0.78, 1.12) |
| Q3 | 0.86 (0.79, 0.95) | 0.80 (0.73, 0.88) | 0.83 (0.68, 1.03) | 0.84 (0.68, 1.04) |
| Q4 | 0.84 (0.75, 0.93) | 0.75 (0.67, 0.85) | 0.79 (0.61, 1.03) | 0.80 (0.62, 1.04) |
| Oesophagus | | | | |
| Q1 | Ref. | Ref. | Ref. | Ref. |
| Q2 | 0.83 (0.73, 0.95) | 0.75 (0.65, 0.86) | 0.99 (0.72, 1.37) | 0.95 (0.69, 1.32) |
| Q3 | 0.69 (0.58, 0.82) | 0.60 (0.50, 0.72) | 0.72 (0.48, 1.08) | 0.68 (0.45, 1.03) |
| Q4 | 0.63 (0.51, 0.79) | 0.53 (0.42, 0.66) | 0.66 (0.39, 1.09) | 0.61 (0.36, 1.02) |
| Lung | | | | |
| Q1 | Ref. | Ref. | Ref. | Ref. |
| Q2 | 0.74 (0.68, 0.81) | 0.74 (0.68, 0.80) | 0.79 (0.65, 0.95) | 0.81 (0.67, 0.98) |
| Q3 | 0.62 (0.56, 0.70) | 0.62 (0.55, 0.69) | 0.70 (0.55, 0.89) | 0.73 (0.56, 0.93) |
| Q4 | 0.50 (0.43, 0.58) | 0.49 (0.42, 0.57) | 0.48 (0.34, 0.66) | 0.50 (0.36, 0.70) |
| Stomach | | | | |
| Q1 | Ref. | Ref. | Ref. | Ref. |
| Q2 | 0.87 (0.76, 0.99) | 0.79 (0.70, 0.90) | 0.94 (0.70, 1.25) | 0.90 (0.67, 1.21) |
| Q3 | 0.83 (0.71, 0.97) | 0.74 (0.63, 0.86) | 0.73 (0.52, 1.02) | 0.69 (0.49, 0.98) |
| Q4 | 0.83 (0.69, 1.00) | 0.71 (0.58, 0.86) | 0.98 (0.65, 1.47) | 0.90 (0.60, 1.37) |
| Pancreas | | | | |
| Q1 | Ref. | Ref. | Ref. | Ref. |
| Q2 | 0.92 (0.83, 1.02) | 0.86 (0.78, 0.95) | 1.11 (0.88, 1.41) | 1.05 (0.83, 1.33) |
| Q3 | 0.92 (0.81, 1.04) | 0.84 (0.74, 0.95) | 1.07 (0.81, 1.41) | 0.98 (0.74, 1.30) |
| Q4 | 0.82 (0.70, 0.96) | 0.73 (0.62, 0.85) | 0.93 (0.66, 1.30) | 0.83 (0.59, 1.18) |
| Liver, bile ducts, and gallbladder | | | | |
| Q1 | Ref. | Ref. | Ref. | Ref. |
| Q2 | 0.83 (0.76, 0.92) | 0.77 (0.69, 0.85) | 1.16 (0.92, 1.47) | 1.10 (0.86, 1.39) |
| Q3 | 0.67 (0.59, 0.76) | 0.60 (0.52, 0.68) | 1.06 (0.80, 1.39) | 0.97 (0.73, 1.28) |
| Q4 | 0.68 (0.58, 0.80) | 0.59 (0.50, 0.69) | 0.87 (0.61, 1.25) | 0.78 (0.54, 1.13) |
| Colon | | | | |
| Q1 | Ref. | Ref. | Ref. | Ref. |
| Q2 | 0.93 (0.87, 1.00) | 0.91 (0.85, 0.97) | 0.90 (0.78, 1.05) | 0.89 (0.76, 1.04) |
| Q3 | 0.88 (0.81, 0.95) | 0.84 (0.78, 0.91) | 0.94 (0.78, 1.13) | 0.92 (0.76, 1.11) |
| Q4 | 0.80 (0.72, 0.88) | 0.75 (0.68, 0.83) | 0.81 (0.64, 1.01) | 0.78 (0.62, 0.99) |
| Rectum | | | | |
| Q1 | Ref. | Ref. | Ref. | Ref. |
| Q2 | 0.96 (0.89, 1.04) | 0.94 (0.87, 1.02) | 0.94 (0.78, 1.12) | 0.90 (0.75, 1.07) |
| Q3 | 0.90 (0.82, 0.99) | 0.88 (0.80, 0.96) | 0.90 (0.73, 1.10) | 0.84 (0.68, 1.04) |
| Q4 | 0.84 (0.75, 0.94) | 0.81 (0.72, 0.91) | 0.75 (0.58, 0.97) | 0.69 (0.53, 0.90) |
| Kidney | | | | |
| Q1 | Ref. | Ref. | Ref. | Ref. |
| Q2 | 0.93 (0.85, 1.02) | 0.84 (0.76, 0.92) | 1.06 (0.86, 1.32) | 1.01 (0.81, 1.26) |
| Q3 | 0.85 (0.76, 0.95) | 0.73 (0.65, 0.82) | 0.98 (0.76, 1.25) | 0.91 (0.70, 1.18) |
| Q4 | 0.79 (0.69, 0.91) | 0.65 (0.57, 0.75) | 0.95 (0.70, 1.29) | 0.86 (0.63, 1.18) |
| Prostate | | | | |
| Q1 | Ref. | Ref. | Ref. | Ref. |
| Q2 | 1.04 (1.01, 1.07) | 1.04 (1.01, 1.07) | 1.00 (0.93, 1.07) | 1.00 (0.94, 1.08) |
| Q3 | 1.05 (1.01, 1.09) | 1.05 (1.01, 1.09) | 1.02 (0.94, 1.11) | 1.03 (0.94, 1.12) |
| Q4 | 1.10 (1.05, 1.15) | 1.10 (1.05, 1.16) | 1.00 (0.90, 1.12) | 1.01 (0.90, 1.13) |
| Bladder | | | | |
| Q1 | Ref. | Ref. | Ref. | Ref. |
| Q2 | 0.92 (0.85, 1.00) | 0.90 (0.82, 0.97) | 0.99 (0.82, 1.20) | 0.98 (0.81, 1.19) |
| Q3 | 0.93 (0.84, 1.02) | 0.89 (0.81, 0.99) | 0.86 (0.69, 1.07) | 0.85 (0.68, 1.05) |
| Q4 | 0.78 (0.69, 0.89) | 0.75 (0.65, 0.85) | 0.91 (0.68, 1.21) | 0.89 (0.66, 1.19) |
| Myeloma | | | | |

| | | | | |
|---|---|---|---|---|
| Q1 | Ref. | Ref. | Ref. | Ref. |
| Q2 | 1.04 (0.90, 1.18) | 0.99 (0.86, 1.13) | 0.78 (0.57, 1.07) | 0.76 (0.55, 1.04) |
| Q3 | 1.07 (0.92, 1.25) | 1.00 (0.86, 1.17) | 0.87 (0.61, 1.26) | 0.83 (0.57, 1.20) |
| Q4 | 1.09 (0.91, 1.31) | 1.00 (0.83, 1.20) | 0.86 (0.55, 1.36) | 0.81 (0.50, 1.29) |
| Melanoma skin | | | | |
| Q1 | Ref. | Ref. | Ref. | Ref. |
| Q2 | 1.17 (1.10, 1.23) | 1.13 (1.07, 1.19) | 1.11 (0.98, 1.25) | 1.08 (0.96, 1.22) |
| Q3 | 1.34 (1.27, 1.43) | 1.28 (1.21, 1.37) | 1.26 (1.11, 1.44) | 1.22 (1.06, 1.40) |
| Q4 | 1.60 (1.50, 1.71) | 1.50 (1.41, 1.61) | 1.36 (1.17, 1.59) | 1.30 (1.11, 1.52) |
| Non-melanoma skin | | | | |
| Q1 | Ref. | Ref. | Ref. | Ref. |
| Q2 | 1.10 (1.06, 1.13) | 1.13 (1.09, 1.17) | 1.01 (0.95, 1.09) | 1.03 (0.96, 1.11) |
| Q3 | 1.17 (1.13, 1.21) | 1.22 (1.18, 1.27) | 1.06 (0.98, 1.15) | 1.09 (1.01, 1.19) |
| Q4 | 1.35 (1.30, 1.40) | 1.44 (1.37, 1.50) | 1.05 (0.96, 1.15) | 1.09 (0.99, 1.20) |

BMI = body mass index. CI = confidence interval. HR = hazard ratio. Q = quartile. All estimates were adjusted for birth cohort, year of conscription, parental education, and parental income. In both cohorts, the median (range) of $W_{max}$ in Q1 was 217 (100-236), in Q2 it was 253 (237-270), in Q3 it was 290 (271-312), in Q4 it was 339 (313-999).

**Supplementary table 8. Net risk and crude risk of cancer at 65 years of age by quartiles of cardiorespiratory fitness in cohort and sibling analysis.**

| | Cohort analysis (N=1 124 049)[a] | | | | Sibling analysis (N=477 453)[a] | | | |
|---|---|---|---|---|---|---|---|---|
| | Net risk[b] | | Crude risk[c] | | Net risk[b] | | Crude risk[c] | |
| Cancer outcome by quartiles of fitness | Risk at age 65 y, % (95% CI) | Difference, pp (95% CI) | Risk at age 65 y, % (95% CI) | Difference, pp (95% CI) | Risk at age 65 y, % (95% CI) | Difference, pp (95% CI) | Risk at age 65 y, % (95% CI) | Difference, pp (95% CI) |
| **Overall cancer diagnosis** | | | | | | | | |
| Q1 | 18.1 (17.8, 18.3) | Ref. | 16.9 (16.7, 17.1) | Ref. | 17.8 (17.3, 18.4) | Ref. | 16.9 (16.4, 17.4) | Ref. |
| Q2 | 18.6 (18.4, 18.9) | 0.6 (0.2, 0.9) | 17.9 (17.7, 18.2) | 1.1 (0.7, 1.5) | 17.8 (17.3, 18.3) | -0.004 (-0.9, 0.8) | 17.2 (16.7, 17.7) | 0.3 (-0.5, 1.1) |
| Q3 | 19.0 (18.6, 19.2) | 0.9 (0.5, 1.3) | 18.3 (18.0, 18.6) | 1.5 (1.1, 1.9) | 18.1 (17.5, 18.6) | 0.2 (-0.7, 1.2) | 17.5 (16.9, 18.0) | 0.6 (-0.3, 1.5) |
| Q4 | 19.5 (19.2, 19.9) | 1.5 (1.0, 1.9) | 19.0 (18.7, 19.4) | 2.2 (1.7, 2.6) | 17.9 (17.3, 18.6) | 0.08 (-0.9, 1.1) | 17.4 (16.7, 18.0) | 0.5 (-0.5, 1.5) |
| **Overall cancer mortality** | | | | | | | | |
| Q1 | 3.0 (2.9, 3.1) | Ref. | 2.8 (2.7, 2.9) | Ref. | 2.8 (2.6, 3.0) | Ref. | 2.6 (2.5, 2.8) | Ref. |
| Q2 | 2.2 (2.1, 2.3) | -0.8 (-0.9, -0.7) | 2.1 (2.0, 2.2) | -0.7 (-0.8, -0.6) | 2.3 (2.1, 2.4) | -0.5 (-0.8, -0.2) | 2.2 (2.0, 2.3) | -0.5 (-0.7, -0.2) |
| Q3 | 2.0 (2.0, 2.1) | -1.0 (-1.1, -0.8) | 2.0 (1.9, 2.1) | -0.8 (-1.0, -0.7) | 2.2 (2.0, 2.4) | -0.6 (-0.9, -0.3) | 2.1 (1.9, 2.3) | -0.5 (-0.8, 0.2) |
| Q4 | 1.9 (1.8, 2.0) | -1.1 (-1.2, 0.9) | 1.9 (1.8, 2.0) | -0.9 (1.1, -0.8) | 2.1 (1.8, 2.3) | -0.7 (-1.1, -0.4) | 2.0 (1.8, 2.2) | -0.7 (-1.0, -0.3) |
| **Site-specific cancers** | | | | | | | | |
| Head and neck | | | | | | | | |
| Q1 | 0.8 (0.7, 0.8) | Ref. | 0.7 (0.7, 0.8) | Ref. | 0.7 (0.6, 0.8) | Ref. | 0.7 (0.6, 0.8) | Ref. |
| Q2 | 0.6 (0.6, 0.6) | -0.2 (-0.3, -0.1) | 0.6 (0.5, 0.6) | -0.2 (-0.2, -0.09) | 0.6 (0.5, 0.7) | -0.1 (-0.3, 0.06) | 0.6 (0.5, 0.7) | -0.1 (-0.2, -0.06) |
| Q3 | 0.6 (0.5, 0.6) | -0.2 (-0.3, -0.1) | 0.5 (0.5, 0.6) | -0.2 (-0.3, -0.1) | 0.6 (0.5, 0.7) | -0.2 (-0.3, -0.02) | 0.5 (0.5, 0.6) | -0.1 (-0.3, 0.03) |
| Q4 | 0.5 (0.5, 0.6) | -0.2 (-0.3, -0.2) | 0.5 (0.5, 0.6) | -0.2 (0.3, -0.1) | 0.5 (0.4, 0.7) | -0.2 (-0.4, -0.02) | 0.5 (0.4, 0.7) | -0.2 (-0.3, 0.03) |
| Oesophagus | | | | | | | | |
| Q1 | 0.3 (0.2, 0.3) | Ref. | 0.3 (0.2, 0.3) | Ref. | 0.2 (0.2, 0.3) | Ref. | 0.2 (0.2, 0.3) | Ref. |
| Q2 | 0.2 (0.1, 0.2) | -0.1 (-0.2, -0.08) | 0.2 (0.1, 0.2) | -0.1 (-0.1, -0.06) | 0.2 (0.1, 0.2) | -0.05 (-0.1, 0.04) | 0.2 (0.1, 0.2) | -0.04 (-0.1, 0.04) |
| Q3 | 0.1 (0.1, 0.2) | -0.1 (-0.2, -0.09) | 0.1 (0.1, 0.2) | -0.1 (-0.2, -0.08) | 0.1 (0.1, 0.2) | -0.08 (-0.2, 0.001) | 0.1 (0.1, 0.2) | -0.07 (-0.2, 0.005) |
| Q4 | 0.1 (0.1, 0.2) | -0.2 (-0.2, -0.1) | 0.1 (0.1, 0.2) | -0.1 (-0.2, -0.09) | 0.1 (0.1, 0.2) | -0.09 (-0.2, -0.004) | 0.1 (0.1, 0.2) | -0.08 (-0.2, 0.001) |
| Lung | | | | | | | | |
| Q1 | 0.6 (0.6, 0.7) | Ref. | 0.6 (0.5, 0.6) | Ref. | 0.6 (0.5, 0.7) | Ref. | 0.5 (0.5, 0.6) | Ref. |
| Q2 | 0.4 (0.3, 0.4) | -0.3 (-0.3, -0.2) | 0.4 (0.3, 0.4) | -0.2 (0.3, -0.2) | 0.4 (0.3, 0.5) | -0.2 (-0.3, -0.07) | 0.4 (0.3, 0.4) | -0.2 (-0.3, -0.06) |
| Q3 | 0.3 (0.3, 0.4) | -0.3 (-0.4, -0.3) | 0.3 (0.3, 0.3) | -0.3 (-0.3, -0.2) | 0.4 (0.3, 0.4) | -0.2 (-0.4, -0.09) | 0.3 (0.3, 0.4) | -0.2 (-0.3, -0.08) |
| Q4 | 0.3 (0.2, 0.3) | -0.4 (-0.4, -0.3) | 0.3 (0.2, 0.3) | -0.3 (-0.4, -0.3) | 0.3 (0.2, 0.3) | -0.3 (-0.5, -0.2) | 0.3 (0.2, 0.3) | -0.3 (-0.4, -0.2) |
| Stomach | | | | | | | | |
| Q1 | 0.3 (0.2, 0.3) | Ref. | 0.3 (0.2, 0.3) | Ref. | 0.3 (0.2, 0.3) | Ref. | 0.2 (0.2, 0.3) | Ref. |
| Q2 | 0.2 (0.2, 0.2) | -0.08 (-0.1, -0.05) | 0.2 (0.2, 0.2) | -0.07 (-0.1, -0.04) | 0.2 (0.2, 0.3) | -0.05 (-0.1, 0.04) | 0.2 (0.2, 0.3) | -0.04 (-0.1, 0.04) |
| Q3 | 0.2 (0.2, 0.2) | -0.09 (-0.1, -0.05) | 0.2 (0.1, 0.2) | -0.08 (-0.1, -0.04) | 0.2 (0.1, 0.2) | -0.09 (-0.2, 0.002) | 0.2 (0.1, 0.2) | -0.08 (-0.2, 0.007) |
| Q4 | 0.2 (0.1, 0.2) | -0.1 (-0.1, -0.06) | 0.2 (0.1, 0.2) | -0.08 (-0.1, -0.04) | 0.2 (0.2, 0.3) | -0.05 (0.2, 0.06) | 0.2 (0.1, 0.3) | -0.04 (-0.1, 0.07) |
| Pancreas | | | | | | | | |
| Q1 | 0.5 (0.4, 0.5) | Ref. | 0.4 (0.4, 0.5) | Ref. | 0.4 (0.3, 0.5) | Ref. | 0.4 (0.3, 0.5) | Ref. |
| Q2 | 0.4 (0.3, 0.4) | -0.1 (-0.2, -0.05) | 0.3 (0.3, 0.4) | -0.1 (-0.1, -0.04) | 0.4 (0.3, 0.5) | 0.0002 (-0.1, 0.1) | 0.4 (0.3, 0.5) | 0.008 (-0.1, 0.1) |
| Q3 | 0.4 (0.3, 0.4) | -0.1 (-0.2, -0.05) | 0.3 (0.3, 0.4) | -0.1 (-0.1, -0.03) | 0.4 (0.3, 0.5) | -0.02 (-0.2, 0.1) | 0.4 (0.3, 0.5) | -0.009 (-0.2, 0.1) |
| Q4 | 0.3 (0.3, 0.4) | -0.2 (-0.2, -0.08) | 0.3 (0.3, 0.4) | -0.1 (-0.2, -0.06) | 0.4 (0.3, 0.5) | -0.07 (-0.2, 0.09) | 0.3 (0.3, 0.5) | -0.05 (-0.2, 0.09) |
| Liver, bile ducts, and gallbladder | | | | | | | | |
| Q1 | 0.5 (0.5, 0.6) | Ref. | 0.5 (0.4, 0.5) | Ref. | 0.4 (0.3, 0.5) | Ref. | 0.3 (0.3, 0.4) | Ref. |
| Q2 | 0.3 (0.3, 0.3) | -0.2 (-0.3, -0.1) | 0.3 (0.3, 0.3) | -0.2 (-0.2, -0.1) | 0.4 (0.3, 0.5) | 0.01 (-0.1, 0.1) | 0.4 (0.3, 0.4) | 0.02 (-0.09, 0.1) |
| Q3 | 0.3 (0.2, 0.3) | -0.3 (-0.3, -0.2) | 0.2 (0.2, 0.3) | -0.2 (-0.3, -0.2) | 0.4 (0.3, 0.4) | -0.02 (-0.2, 0.1) | 0.3 (0.3, 0.4) | -0.01 (-0.1, 0.01) |
| Q4 | 0.3 (0.2, 0.3) | -0.3 (-0.3, -0.2) | 0.2 (0.2, 0.3) | -0.2 (-0.3, -0.2) | 0.3 (0.2, 0.4) | -0.08 (-0.2, 0.06) | 0.3 (0.2, 0.4) | -0.06 (-0.2, 0.07) |
| Colon | | | | | | | | |

| | | | | | | | | |
|---|---|---|---|---|---|---|---|---|
| Q1 | 1.2 (1.1, 1.2) | Ref. | 1.1 (1.0, 1.1) | Ref. | 1.1 (1.0, 1.3) | Ref. | 1.0 (0.9, 1.2) | Ref. |
| Q2 | 0.9 (0.9, 1.0) | -0.2 (-0.3, -0.1) | 0.9 (0.8, 1.0) | -0.2 (-0.3, -0.09) | 0.9 (0.8, 1.0) | -0.2 (-0.4, -0.02) | 0.9 (0.7, 1.0) | -0.2 (-0.4, 0.03) |
| Q3 | 0.9 (0.8, 1.0) | -0.3 (-0.4, -0.2) | 0.9 (0.8, 0.9) | -0.2 (-0.3, -0.1) | 0.9 (0.8, 1.1) | -0.2 (-0.4, 0.07) | 0.9 (0.8, 1.0) | -0.1 (-0.4, 0.09) |
| Q4 | 0.8 (0.8, 0.9) | -0.3 (-0.5, -0.2) | 0.8 (0.7, 0.9) | -0.3 (-0.4, -0.2) | 0.8 (0.7, 1.0) | -0.3 (-0.5, -0.02) | 0.8 (0.7, 0.9) | -0.2 (-0.5, 0.008) |
| Rectum | | | | | | | | |
| Q1 | 0.9 (0.8, 0.9) | Ref. | 0.8 (0.7, 0.9) | Ref. | 0.9 (0.7, 1.0) | Ref. | 0.8 (0.7, 0.9) | Ref. |
| Q2 | 0.7 (0.7, 0.8) | -0.1 (-0.2, -0.04) | 0.7 (0.7, 0.8) | -0.1 (-0.2, -0.007) | 0.7 (0.6, 0.8) | -0.2 (-0.4, 0.003) | 0.6 (0.5, 0.8) | -0.2 (-0.4, 0.02) |
| Q3 | 0.7 (0.6, 0.8) | -0.2 (-0.3, -0.07) | 0.7 (0.6, 0.7) | -0.1 (-0.2 (-0.03) | 0.6 (0.5, 0.8) | -0.2 (-0.4, 0.01) | 0.6 (0.5, 0.7) | -0.2 (-0.4, 0.007) |
| Q4 | 0.7 (0.6, 0.7) | -0.2 (-0.3, -0.09) | 0.6 (0.6, 0.7) | -0.2 (-0.3, -0.05) | 0.6 (0.5, 0.7) | -0.3 (-0.5, -0.09) | 0.5 (0.4, 0.7) | -0.3 (-0.5, -0.007) |
| Kidney | | | | | | | | |
| Q1 | 0.6 (0.6, 0.7) | Ref. | 0.6 (0.5, 0.6) | Ref. | 0.5 (0.4, 0.6) | Ref. | 0.5 (0.4, 0.6) | Ref. |
| Q2 | 0.4 (0.4, 0.5) | -0.2 (-0.3, -0.1) | 0.4 (0.4, 0.5) | -0.2 (-0.2, -0.1) | 0.5 (0.4, 0.6) | -0.03 (-0.2, 0.1) | 0.5 (0.4, 0.6) | -0.01 (-0.2, 0.1) |
| Q3 | 0.4 (0.4, 0.5) | -0.2 (-0.3, -0.2) | 0.4 (0.3, 0.4) | -0.2 (-0.3, -0.1) | 0.5 (0.4, 0.6) | -0.06 (-0.2, 0.1) | 0.4 (0.4, 0.5) | -0.05 (-0.2, 0.1) |
| Q4 | 0.4 (0.3, 0.4) | -0.3 (-0.4, -0.2) | 0.4 (0.3, 0.4) | -0.2 (-0.3, -0.2) | 0.4 (0.3, 0.6) | -0.08 (-0.3, 0.09) | 0.4 (0.3, 0.5) | -0.06 (-0.2, 0.1) |
| Prostate | | | | | | | | |
| Q1 | 4.9 (4.7, 5.1) | Ref. | 4.5 (4.3, 4.7) | Ref. | 5.0 (4.7, 5.4) | Ref. | 4.7 (4.3, 5.0) | Ref. |
| Q2 | 5.3 (5.1, 5.5) | 0.4 (0.2, 0.6) | 5.0 (4.8, 5.2) | 0.5 (0.3, 0.7) | 5.1 (4.8, 5.5) | 0.06 (-0.4, 0.6) | 4.8 (4.5, 5.2) | 0.2 (-0.3, 0.6) |
| Q3 | 5.4 (5.1, 5.6) | 0.4 (0.2, 0.7) | 5.1 (4.9, 5.3) | 0.6 (0.4, 0.9) | 5.2 (4.8, 5.6) | 0.2 (-0.4, 0.7) | 4.9 (4.5, 5.3) | 0.3 (-0.3, 0.8) |
| Q4 | 5.5 (5.3, 5.8) | 0.6 (0.3, 0.9) | 5.3 (5.1, 5.6) | 0.8 (0.6, 1.1) | 5.1 (4.7, 5.6) | 0.08 (-0.6, 0.8) | 4.9 (4.5, 5.4) | 0.2 (-0.4, 0.9) |
| Bladder | | | | | | | | |
| Q1 | 0.7 (0.6, 0.7) | Ref. | 0.6 (0.6, 0.7) | Ref. | 0.6 (0.5, 0.7) | Ref. | 0.6 (0.5, 0.6) | Ref. |
| Q2 | 0.5 (0.5, 0.6) | -0.1 (-0.2, -0.06) | 0.5 (0.5, 0.6) | -0.1 (-0.2, -0.03) | 0.5 (0.5, 0.6) | -0.05 (-0.2, 0.09) | 0.5 (0.4, 0.6) | -0.04 (-0.2, 0.09) |
| Q3 | 0.5 (0.5, 0.6) | -0.1 (-0.2, -0.05) | 0.5 (0.5, 0.6) | -0.1 (-0.2, -0.02) | 0.5 (0.4, 0.6) | -0.1 (-0.3, 0.04) | 0.5 (0.4, 0.6) | -0.09 (-0.2, 0.05) |
| Q4 | 0.5 (0.4, 0.5) | -0.2 (-0.3, -0.1) | 0.5 (0.4, 0.5) | -0.2 (-0.2, -0.08) | 0.5 (0.4, 0.6) | -0.09 (-0.3, 0.09) | 0.5 (0.4, 0.6) | -0.07 (-0.2, 0.1) |
| Myeloma | | | | | | | | |
| Q1 | 0.3 (0.2, 0.3) | Ref. | 0.2 (0.2, 0.3) | Ref. | 0.3 (0.2, 0.4) | Ref. | 0.3 (0.2, 0.4) | Ref. |
| Q2 | 0.3 (0.2, 0.3) | -0.004 (-0.05, 0.04) | 0.3 (0.2, 0.3) | 0.006 (-0.04, 0.005) | 0.2 (0.2, 0.3) | -0.09 (-0.2, 0.02) | 0.2 (0.2, 0.3) | -0.09 (-0.2, 0.02) |
| Q3 | 0.3 (0.2, 0.3) | 0.00001 (-0.05, 0.05) | 0.3 (0.2, 0.3) | 0.01 (-0.04, 0.06) | 0.2 (0.2, 0.3) | -0.08 (-0.2, 0.05) | 0.2 (0.2, 0.3) | -0.07 (-0.2, 0.05) |
| Q4 | 0.3 (0.2, 0.3) | 0.001 (-0.06, 0.06) | 0.3 (0.2, 0.3) | 0.01 (-0.04, 0.07) | 0.2 (0.2, 0.3) | -0.08 (-0.2, 0.06) | 0.2 (0.1, 0.3) | -0.07 (-0.2, 0.06) |
| Melanoma skin | | | | | | | | |
| Q1 | 1.7 (1.5, 1.7) | Ref. | 1.5 (1.4, 1.6) | Ref. | 1.7 (1.5, 1.8) | Ref. | 1.5 (1.4, 1.7) | Ref. |
| Q2 | 2.2 (2.1, 2.3) | 0.6 (0.5, 0.8) | 2.1 (2.0, 2.2) | 0.6 (0.5, 0.8) | 2.1 (1.8, 2.3) | 0.4 (0.06, 0.7) | 2.0 (1.8, 2.2) | 0.5 (0.09, 0.7) |
| Q3 | 2.4 (2.3, 2.6) | 0.8 (0.6, 1.0) | 2.3 (2.2, 2.5) | 0.9 (0.7, 1.0) | 2.2 (2.0, 2.5) | 0.6 (0.2, 0.9) | 2.1 (1.9, 2.4) | 0.6 (0.2, 1.0) |
| Q4 | 2.7 (2.5, 2.8) | 1.0 (0.9, 1.3) | 2.6 (2.4, 2.7) | 1.1 (0.9, 1.3) | 2.3 (2.0, 2.6) | 0.6 (0.2, 1.1) | 2.2 (2.0, 2.5) | 0.7 (0.3, 1.1) |
| Non-melanoma skin | | | | | | | | |
| Q1 | 4.4 (4.2, 4.5) | Ref. | 4.0 (3.9, 4.1) | Ref. | 4.7 (4.4, 5.0) | Ref. | 4.4 (4.2, 4.7) | Ref. |
| Q2 | 5.7 (5.6, 5.9) | 1.4 (1.1, 1.6) | 5.5 (5.3, 5.6) | 1.4 (1.2, 1.7) | 5.1 (4.8, 5.4) | 0.4 (-0.1, 0.9) | 4.9 (4.6, 5.2) | 0.5 (-0.02, 0.9) |
| Q3 | 6.1 (5.9, 6.3) | 1.7 (1.4, 2.0) | 5.8 (5.6, 6.0) | 1.8 (1.6, 2.1) | 5.3 (5.0, 5.7) | 0.6 (0.03, 0.1) | 5.1 (4.8, 5.4) | 0.7 (0.1, 1.2) |
| Q4 | 6.7 (6.5, 7.0) | 2.4 (2.1, 2.7) | 6.5 (6.3, 6.7) | 2.5 (2.2, 2.8) | 5.3 (4.9, 5.7) | 0.6 (-0.05, 0.1) | 5.1 (4.7, 5.5) | 0.7 (0.1, 1.2) |

[a]The flexible parametric models were performed in the full sample, from which the standardised incidences were computed in a random subsample of 10%.

[b]Standardised cumulative incidences as obtained in the main analysis, assuming conditional independence between time to the cancer outcome and the competing event (death from other causes).

[c]Cause-specific standardised cumulative incidence functions, accounting for the competing risk of death from non-cancer causes.

CI = confidence interval. Q = quartile. All estimates are adjusted for age at conscription, year of conscription, body mass index, parental education, and parental income. In both cohorts, the median (range) of $W_{max}$ in Q1 was 217 (100-236), in Q2 it was 253 (237-270), in Q3 it was 290 (271-312), in Q4 it was 339 (313-999).

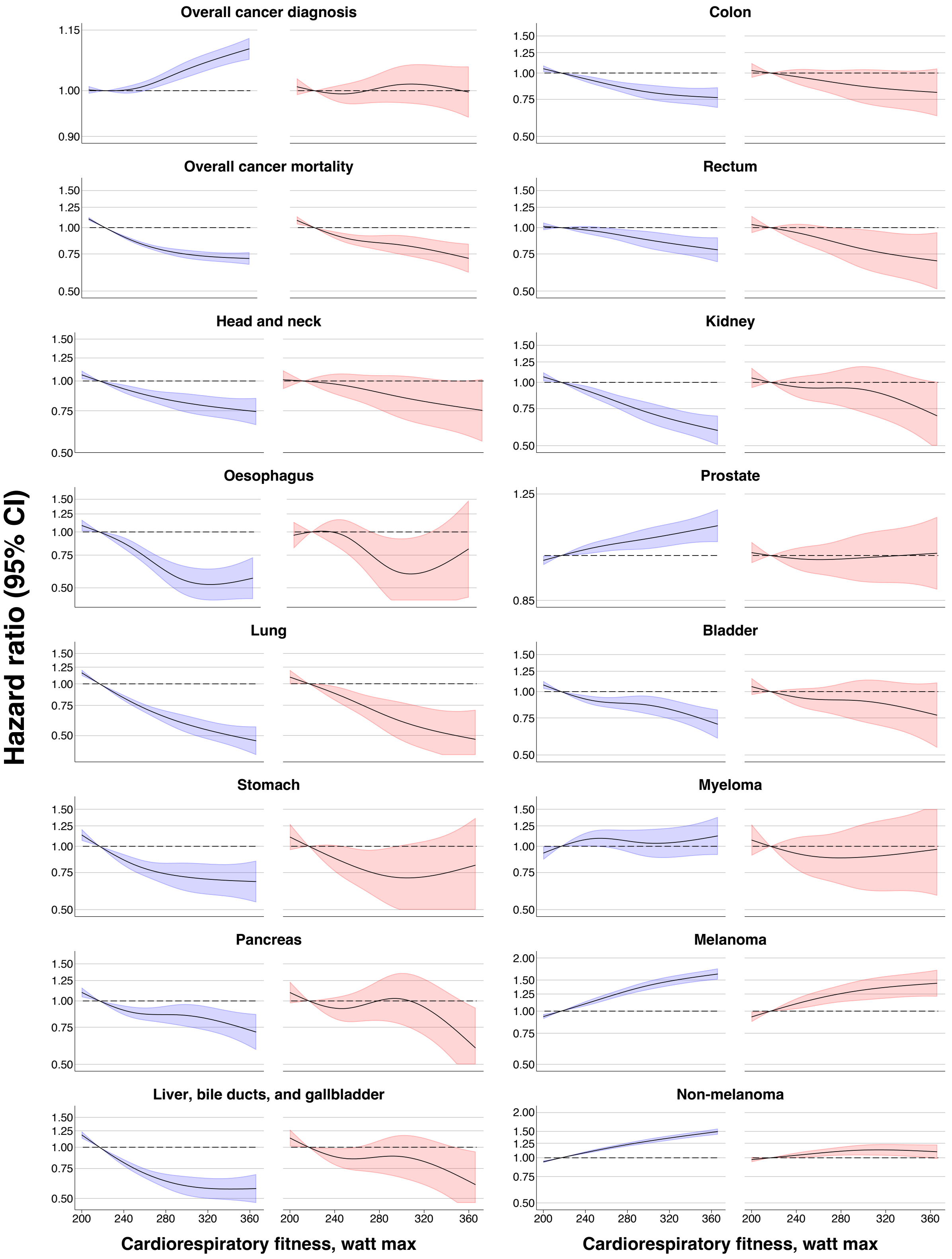


**Supplementary figure 1. Hazard ratios for cancer across restricted cubic splines of cardiorespiratory fitness in cohort (blue) and sibling analysis (red).** Estimates were obtained using flexible parametric survival models, extended to a marginalised between-within model in the sibling cohort, with knots placed at the $5^{th}$, $35^{th}$, $65^{th}$, and $95^{th}$ percentile, and using age as the underlying time scale. The referent was set to the median value of the bottom quartile (217 $W_{max}$). The models were adjusted for age at conscription, year of conscription, body mass index, parental education, and parental income. For graphical purposes, the x-axis was limited to span from the $5^{th}$ to the $95^{th}$ percentile of the distribution.

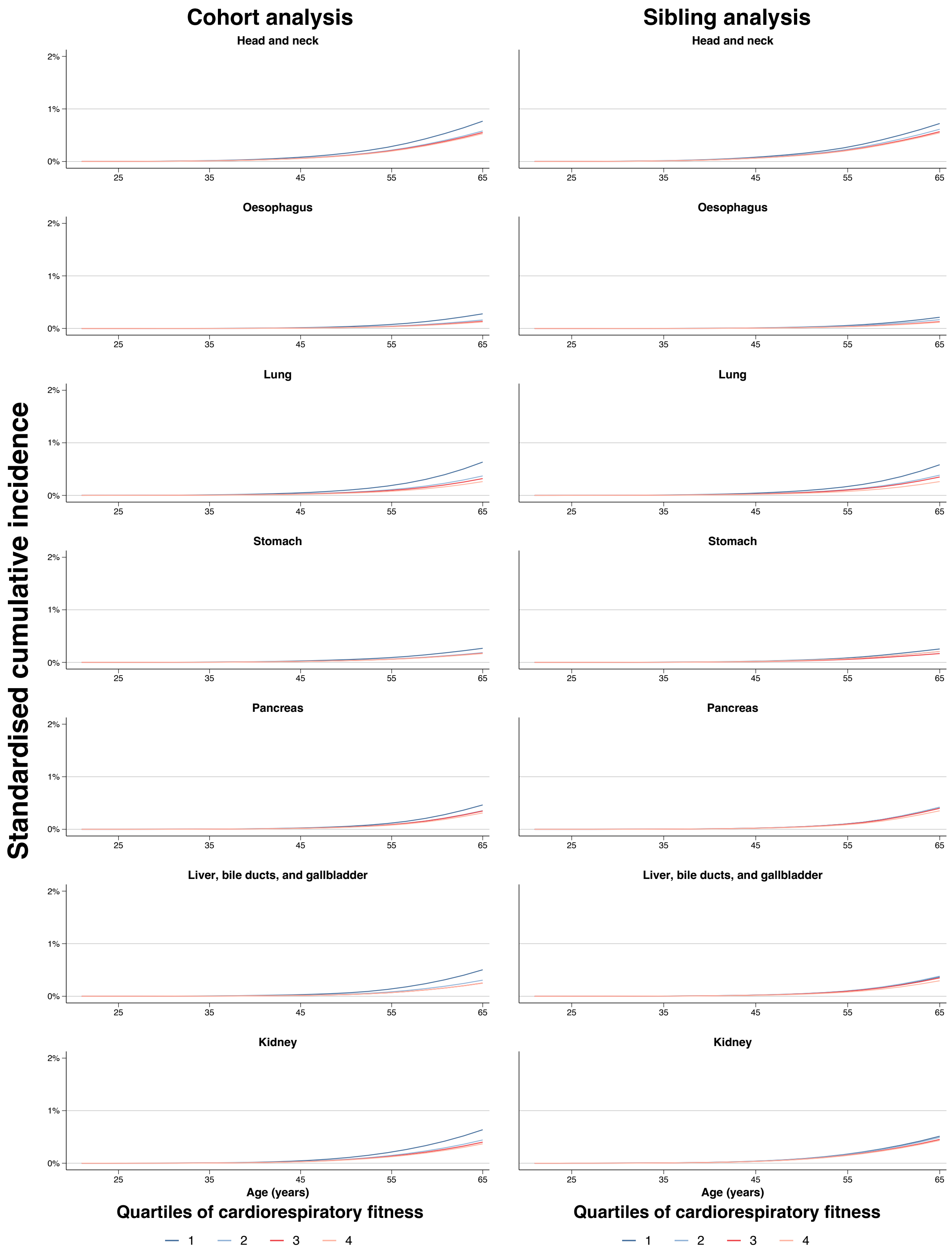


**Supplementary figure 2. Standardised cumulative incidence for site-specific cancer by quartiles of cardiorespiratory fitness in cohort and sibling analysis.** Estimates obtained using flexible parametric survival models, extended to a marginalised between-within model in the sibling cohort, with baseline knots placed at the 5$^{th}$, 27.5$^{th}$, 50$^{th}$, 72.5$^{th}$, and 95$^{th}$ percentile of the uncensored log survival times, and using age as the underlying time scale. All models were adjusted for age at conscription, year of conscription, body mass index, parental education, and parental income. Inferential measures for the incidences were omitted for clarity as they are also reflected in supplementary table 3 and 4.

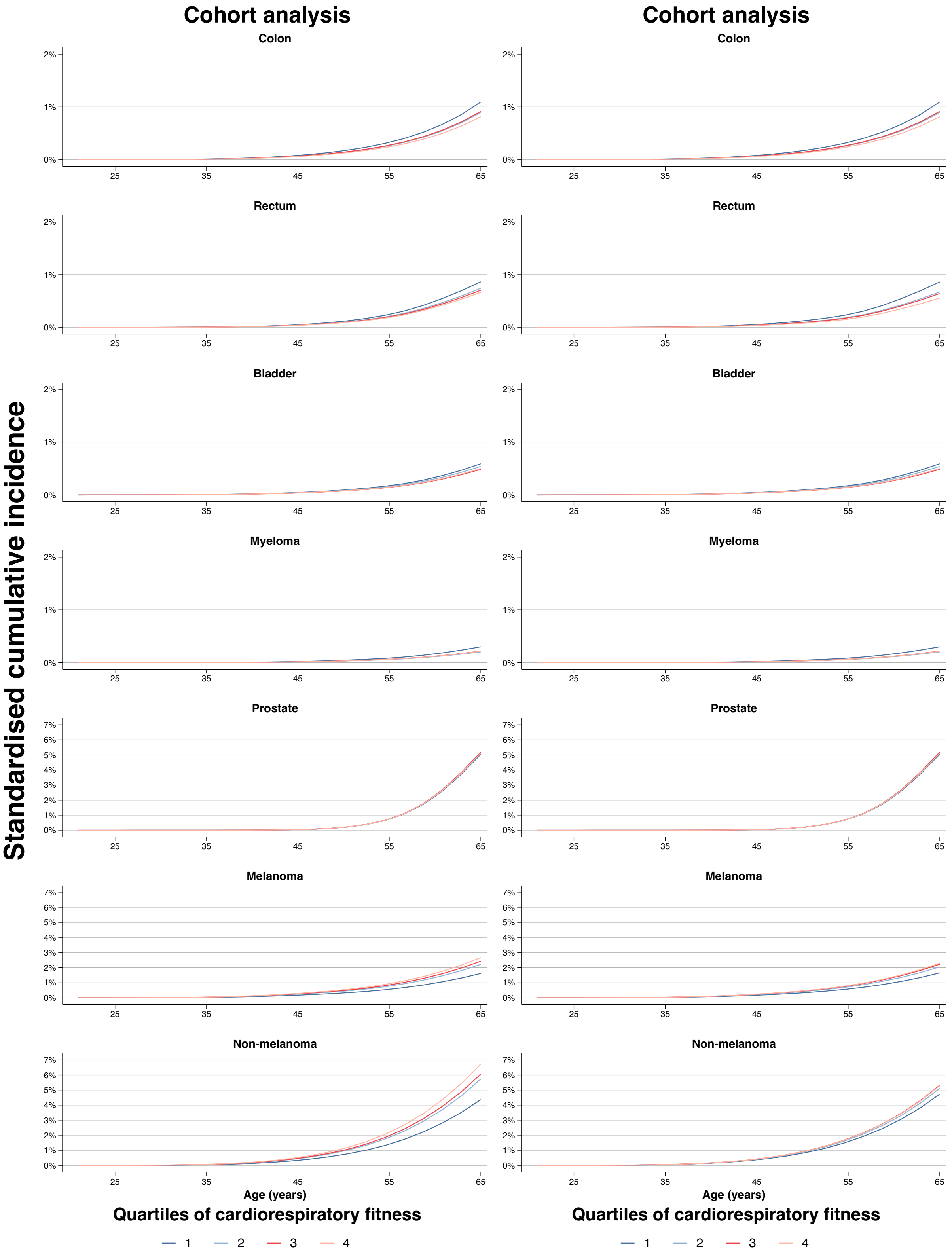


**Supplementary figure 3. Standardised cumulative incidence for site-specific cancer by quartiles of cardiorespiratory fitness in cohort and sibling analysis.** Estimates obtained using flexible parametric survival models, extended to a marginalised between-within model in the sibling cohort, with baseline knots placed at the 5$^{th}$, 27.5$^{th}$, 50$^{th}$, 72.5$^{th}$, and 95$^{th}$ percentile of the uncensored log survival times, and using age as the underlying time scale. All models were adjusted for age at conscription, year of conscription, body mass index, parental education, and parental income. Inferential measures for the incidences were omitted for clarity as they are also reflected in supplementary table 3 and 4.
